# HPV prevalence and associated factors in Cameroon: a systematic review and meta-analysis

**DOI:** 10.64898/2026.02.15.26346335

**Authors:** Fabrice Zobel Lekeumo Cheuyem, Rick Tchamani, Edmond Martial Lemaire Bodo, Chabeja Achangwa, Solange Dabou, Mohamadou Adama, Doh Gilbert Ndeh

**Author notes:** **Corresponding author’s address**: Fabrice Zobel Lekeumo Cheuyem.

## Abstract

**Background:** Cervical cancer, generally induced by human papillomavirus (HPV) infection remains one of the most prevalent and deadly female cancers in sub-Saharan Africa (SSA). In Cameroon, the impact of prevention strategies is limited by systemic challenges, and insufficient evidence base to guide effective interventions. This study aimed to synthesize available evidence on the prevalence and key determinants of HPV infection among Cameroonian women.

**Methods:** A comprehensive search was conducted across PubMed, Scopus, Web of Science, Embase, Cochrane electronic databases and local online publishers. Quality assessment of included studies was performed using the Joanna Briggs Institute (JBI) critical appraisal tool. The random effect model was used to pooled the estimates. Heterogeneity was evaluated using the *I*^2^ statistics. Statistical significance was set at *p* <0.05 and all analyses were conducted using R Statistics version 4.5.2. The protocol was registered on PROSPERO (CRD420261279093).

**Results:** Thirty-six studies (20,033 participants) were included. The pooled prevalence of HPV infection 36.10 (95% CI: 27.28-45.97) with high heterogeneity (*I^2^* = 98.4%). Higher estimates were observed among female sex workers 62.10% (95% CI: 58.08–66.00%, 1 study, n = 599) and women with pre-cancerous genital lesions 85.53% (95% CI: 61.72–95.59%, 4 studies, n = 673). Significant determinants of HPV infection included age below 40 (OR = 1.31; 95% CI: 1.14–1.49; 7 reports), unmarried status (OR = 1.43; 95% CI: 1.24–1.64; 15 reports), having five or more sexual partners (OR = 1.26; 95% CI: 1.05–1.51; 2 reports), parity below four (OR = 1.29; 95% CI: 1.09–1.52; 2 reports), HIV infection (OR = 1.92; 95% CI: 1.24–2.98; 6 reports), CD4 count below 500 cells/mm³ (OR = 2.00; 95% CI: 1.02–3.95; 2 reports), and viral load below 1000 copies/mL (OR = 2.12; 95% CI: 1.27–3.53; 2 reports).

**Conclusions:** Our study demonstrates a high and persistent burden of HPV infection in Cameroon, with a greater impact on younger women and women living with HIV. These findings highlight an urgent public health need to strengthen and expand prevention strategies to effectively reduce and eliminate cervical cancer incidence in the country.

## 1. Introduction

Human papillomavirus (HPV) is a common sexually transmitted infection affecting the mucocutaneous epithelium of the anogenital tract and other anatomical sites [1,2]. Among the more than 170 identified genotypes, at least 15 (16, 18, 31, 33, 35, 39, 45, 51, 52, 56, 58, 59, 68, 73, and 82) are classified as high-risk human papillomavirus (HR-HPV) due to their oncogenic potential and involvement in precancerous cervical lesions [3,4]. In the absence of treatment, these lesions may progress to invasive cervical cancer, a malignant tumor arising from cervical cells [5,6]. Nearly 99% of cervical cancer cases are attributable to persistent HR-HPV infection. Cervical cancer ranks as the fourth most common cancer among women worldwide; in 2022 alone, it caused an estimated 350,000 deaths [7]. The progression from HR-HPV infection to cervical cancer is influenced by several concurrent factors, including immunosuppression, smoking, high parity, long-term oral contraceptive use, and obesity [8].

Cervical cancer remains a major public health concern in Africa. According to estimates from the HPV Information Centre, more than 117,000 new cases of cervical cancer are diagnosed annually across the continent, and over 76,000 deaths are reported each year, making it the second leading causes of cancer-related mortality among women in the region [9]. In sub-Saharan Africa, approximately 110,300 new cases of cervical cancer are reported annually, and women have an estimated 3 to 5% lifetime risk of developing the disease before the age of 75 [10]. HR-HPV genotypes, particularly HPV-16 and HPV-18 have been identified as the principal etiological agents, accounting for more than 65% of cervical cancer cases in SSA [11]. Despite the proven effectiveness of prevention strategies, including HPV vaccination and cervical cancer screening in reducing the disease incidence and mortality, their implementation in SSA remains suboptimal. Although available in the region, vaccine and screening coverage are low, due to persistent barriers related to access, awareness, health system capacity, and resource availability [12–15].

In Cameroon, approximately 2,770 new cases of cervical cancer are diagnosed each year, with an estimated 1,787 deaths reported annually [16]. In 2023, several studies conducted in different settings highlighted substantial variability in HPV prevalence across the country. A hospital-based study carried out in the North-West region reported a prevalence of 36.16% [17]. Besides, a community-based study conducted in the Littoral region found a higher prevalence of 57.89%, while a hospital-based study in the Centre region reported a prevalence of 30.61% [18,19]. In Cameroon like in other sub-Saharan Africa countries, the implementation of cervical cancer prevention measures remains insufficient and inadequate [14,20]. The most vulnerable populations include sex workers and people living with human Immunodeficiency Virus (HIV) [21]. Several factors, including early-age sexual intercourse and childbirth, having multiple sexual partners, being uneducated and using injectable contraception have been identified as predictors of HPV infections. [22,23].

However, despite a growing body of evidence, the overall prevalence of HPV infection and the magnitude of associated risk factors among women in Cameroon have not been systematically synthesized. Given the high burden of HPV and the variability of reported prevalence and risk factors across the country, a comprehensive synthesis of existing evidence is urgently needed to strengthen the evidence base necessary for designing targeted and effective public health interventions and shaping future cervical cancer related health policies in Cameroon. Against this backdrop, the present systematic review and meta-analysis was conducted to estimate the pooled prevalence of HPV infection among Cameroonian women and identify key determinant of HPV infection.

## 2. Methods

### 2.1. Study design

This review was conducted in accordance with the Preferred Reporting Items for Systematic Reviews and Meta-Analyses (PRISMA) guidelines and was registered with the International Prospective Register of Systematic Reviews (CRD420261279093).

### 2.2. Eligibility criteria

#### 2.2.1. Inclusion criteria

This systematic review included observational studies (cross-sectional, cohort, or case-control) that report on the prevalence and/or risk factors of HPV in Cameroon. Interventional studies providing outcomes of interest were also included. The population comprised sexually active individuals at risk of contracting an HPV infection. Eligible studies were restricted to those published in English or French. However, no restrictions regarding the study period were applied.

#### 2.2.2. Exclusion criteria

Studies with duplicated information, with research topics not related to the objective of our study, case reports, posters, conference papers, reviews, editorials, commentaries, and animal-based studies were not considered for this study. In addition, articles without full text were disregarded due to insufficient data. Moreover, studies conducted out of Cameroon were not included.

### 2.3. Search Strategy

To identify all relevant literature for this review, we conducted a comprehensive search across several electronic databases: PubMed, Scopus, Web of Science, Embase, and the Cochrane Library. To ensure the inclusion of research by Cameroonian scholars and local journals, we also searched African Journals Online (AJOL) and the Health Sciences and Disease journal platform. Additionally, we searched for grey literature, including unpublished research and preprints. Manual searches were also performed using Google Scholar and by screening the reference lists of the included studies.

To identify relevant literature effectively, we employed a search strategy combining Medical Subject Headings (MeSH) terms and Boolean operators. The specific search string used in PubMed was as follows: (“human papillomavirus”[tiab] OR HPV[tiab] OR “cervical cancer”[tiab] OR “uterine cervical neoplasms”[Mesh]) AND (Cameroon[tiab] OR “Cameroon”[Mesh]) (**Supplementary Table 1**). Articles retrieved were imported into the Zotero reference manager (version 7.0.32, Corporation for Digital Scholarship, Fairfax, Virginia, USA), and duplicate articles were removed. Screening was conducted by two investigators (FZLC and RT), and any disagreements were resolved through discussion to reach a consensus. The literature search was completed on January 27, 2025.

### 2.4. Data extraction

We extracted key data from each study into a Microsoft Excel spreadsheet. For each study, we recorded the following information: the first author’s last name, the year of study completion, the region of Cameroon where the study was conducted, the type of participants, the sampling method, the diagnostic tool used, the sample size, the number of eligible participants screened for HPV, and the number of positive cases detected. Regarding determinant outcomes, odds ratios and their corresponding 95% confidence intervals (CIs) were also collected. This step was performed by one investigator (FZLC) and cross-checked by two other research team members (ELMB and DGN).

### 2.5. Outcomes measurement

The prevalence was calculated by dividing the number of HPV-positive cases by the total number of participants tested and multiplying by 100. An HPV-positive case was defined as any participant with a valid sample collected either by a healthcare provider or through self-sampling that tested positive via genotyping or an equivalent validated laboratory assay.

### 2.6. Risk of bias assessment

Two independent reviewers (FZLC and CA) assessed study quality and risk of bias using the Joanna Briggs Institute (JBI) critical appraisal tools tailored to each study design. Disagreements were resolved through discussion or by consulting a third reviewer (DGN). For the cross-sectional study the criteria included studies clear definition of inclusion criteria, comprehensive descriptions of study subjects and settings, validity and reliability of exposure measurements, use of objective, standardized criteria for outcome assessment, identification potential confounding factors, implementation of appropriate strategies to address them, and the appropriateness of statistical method. Each criterion was scored as 1 (yes) or 0 (no or unclear). In this study we defined and apply the following categorization of the overall risk of bias: low (>50%), moderate (>25-50%), or high (≤25%).

### 2.7. Formal analysis

We used a random-effects model (DerSimonian-Laird method) to generate the HPV prevalence and determinant as we anticipated a high heterogeneity between studies. The *I^2^* statistics was used to assess heterogeneity between studies. To pooled the HPV prevalence, we used the Generalized Linear Mixed Models (GLMM), coupled with the Probit-Logit Transformation (PLOGIT) because of their effectiveness in handling meta-analyses of binary data [24]. Statistical significance was set at a *p*-value of <0.05. All analyses including assessing correlated of HPV positivity was conducted using the ‘meta’ package in R Statistics version 4.5.2 [25].

### 2.8. Publication bias and sensitivity analysis

We assessed publication bias using funnel plots and formal statistical tests, such as Egger’s or Begg’s tests [26,27]. A *p*-value < 0.05 was considered indicative of statistically significant publication bias. In such cases, the trim-and-fill method was used to adjust the pooled estimate. Additionally, sensitivity analysis was performed by iteratively excluding one study at a time (leave-one-out method) to assess the robustness of the findings.

## 3. Results

The systematic search identified 741 records (733 from databases and 8 from other sources). After removing 43 duplicates, we screened 698 records by title and abstract and then assessed 57 by full-text. Of these, 36 primary research articles met the eligibility criteria and were included in the systematic review and meta-analysis (**Fig. 1**).

**Fig. 1.**
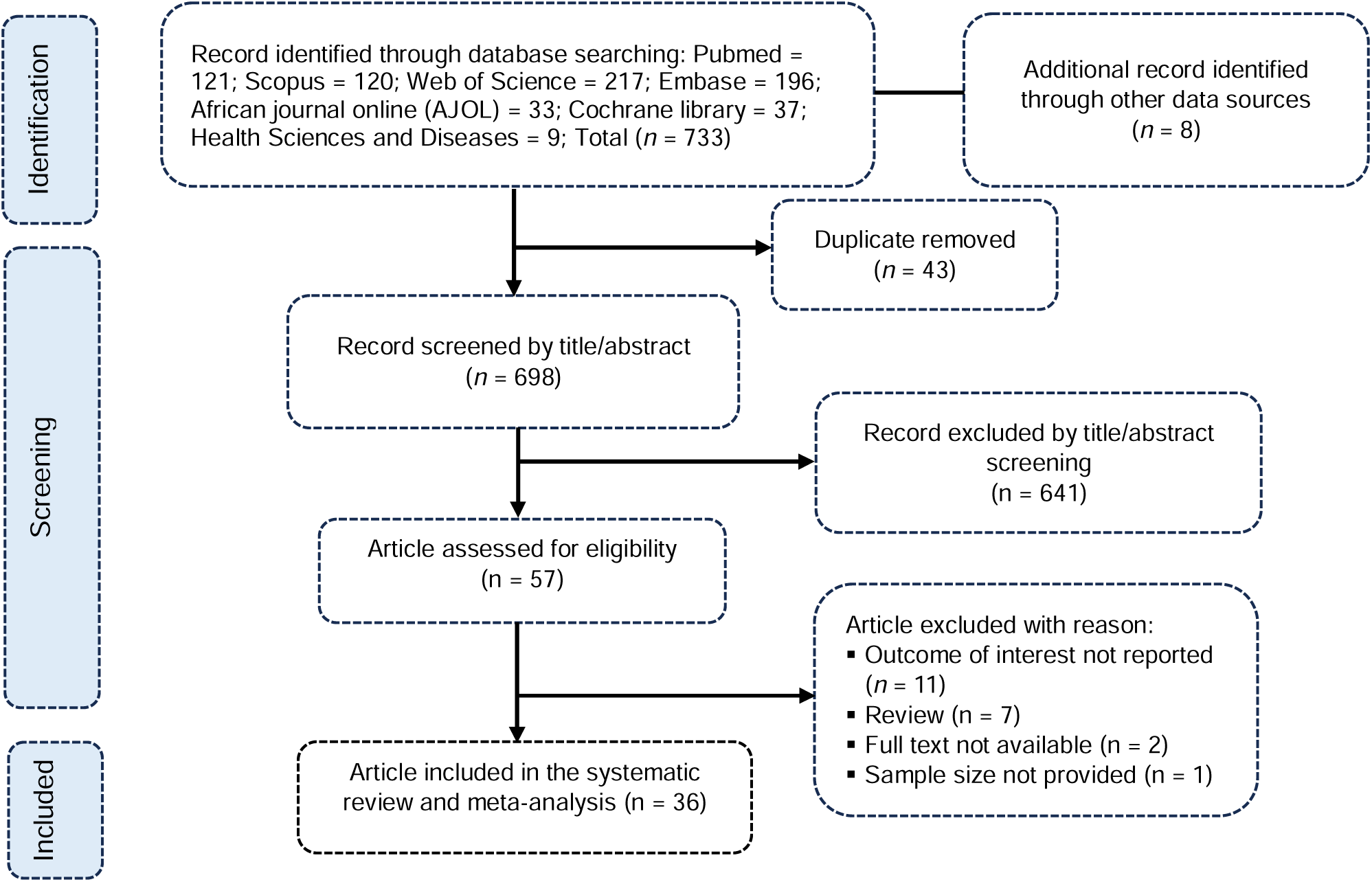
PRISMA diagram flow from study identification to inclusion in the meta-analysis

### 3.1. Studies selection

### 3.2. Study characteristics

The systematic review included 36 studies encompassing a total of 20,033 participants, with the vast majority utilizing a cross-sectional design (34 studies, 94.4%). Research efforts were concentrated geographically, with 10 studies (27.8%) conducted in the Centre region and 9 studies (25.0%) in the West region. The studies covered diverse at-risk populations: 26 studies (72.2%) focused on women from the general population, while specific high-risk groups were also investigated, including 4 studies (11.1%) on people living with HIV (PLHIV) and 4 studies (11.1%) on women with pre-cancerous or cancerous lesions. The settings were predominantly clinical, with 26 studies (72.2%) conducted in hospitals, compared to 9 community-based studies (25.0%). The reported HPV prevalence across the 36 studies showed high variability, ranging from 4.95% to 96.69%. Diagnostic methods were heterogeneous but dominated by PCR-based techniques. The Abbott RealTime High-Risk HPV assay was the most frequently used tool, appearing in 10 studies (27.8%), followed by the Cepheid GeneXpert system in 6 studies (16.7%). Most studies used non-probability sampling methods (34 studies, 94.4%). Furthermore, the risk of bias was rated as low in 34 studies (94.4%) (**Table 1**).

**Table 1.** Determinants of human papillomavirus infection in Cameroon.

| Predictor | Modality | OR<sup>1</sup> | 95% CI |  | Reports included | Heterogeneity |  |
| --- | --- | --- | --- | --- | --- | --- | --- |
|  |  |  | Lower | Upper |  | I<sup>2</sup> (%) | p-value |
| Age (years) | ≤ 40 vs. 40 + years | 1.31 | 1.14 | 1.49 | 7 | 43.4 | 0.101 |
| Educational level | Tertiary vs. Other<sup>2</sup> | 0.93 | 0.80 | 1.08 | 11 | 18.2 | 0.271 |
| Marital status | Unmarried vs. in Partnership | 1.43 | 1.24 | 1.64 | 15 | 0.0 | 0.640 |
| Occupation | Housewife vs. Other | 0.96 | 0.70 | 1.33 | 8 | 40.5 | 0.109 |
| Employment status | Employed vs. Other | 1.02 | 0.78 | 1.33 | 4 | 52.2 | 0.099 |
| Menarche (years) | ≤ 13 vs. 13 + | 1.07 | 0.89 | 1.30 | 2 | 0.0 | 0.810 |
| Coitarche | ≤ 16 vs. 16 + | 1.25 | 0.95 | 1.66 | 7 | 15.4 | 0.313 |
| Number of sexual partners | 2 + vs. ≤ 2 | 1.16 | 0.90 | 1.49 | 6 | 0.0 | 0.691 |
|  | 5 + vs. ≤ 5 | 1.26 | 1.05 | 1.51 | 2 | 0.0 | 0.873 |
| Contraception | Hormonal vs. None | 1.39 | 1.09 | 1.77 | 3 | 31.8 | 0.231 |
|  | Condom vs. None | 1.61 | 1.19 | 2.17 | 4 | 0.0 | 0.510 |
| Parity | ≤ 4 vs. 4 + | 1.29 | 1.09 | 1.52 | 2 | 0.0 | 0.375 |
| HIV status | Positive vs. Negative | 1.92 | 1.24 | 2.98 | 6 | 72.6 | 0.003 |
| CD4 count (cells/mm<sup>3</sup>) | ≤ 500 vs. 500 + | 2.00 | 1.02 | 3.95 | 2 | 0.0 | 0.532 |
| Viral load (copies/mL) | ≤ 1000 vs. 1000 + | 2.12 | 1.27 | 3.53 | 2 | 0.0 | 0.839 |

<sup>1</sup>Random effect model; <sup>2</sup> Other included No formal education, primary or secondary school; Unmarried included single, divorced, widow; In partnership included married or cohabitation

### 3.3. HPV prevalence & subgroup analysis

The pooled prevalence of HPV infection among 20,033 Cameroonians from 36 studies was 36.10 (95% confidence interval (CI): 27.28-45.97) with high heterogeneity (*I^2^* = 98.4%) (**Fig. 2**).

**Fig. 2.**
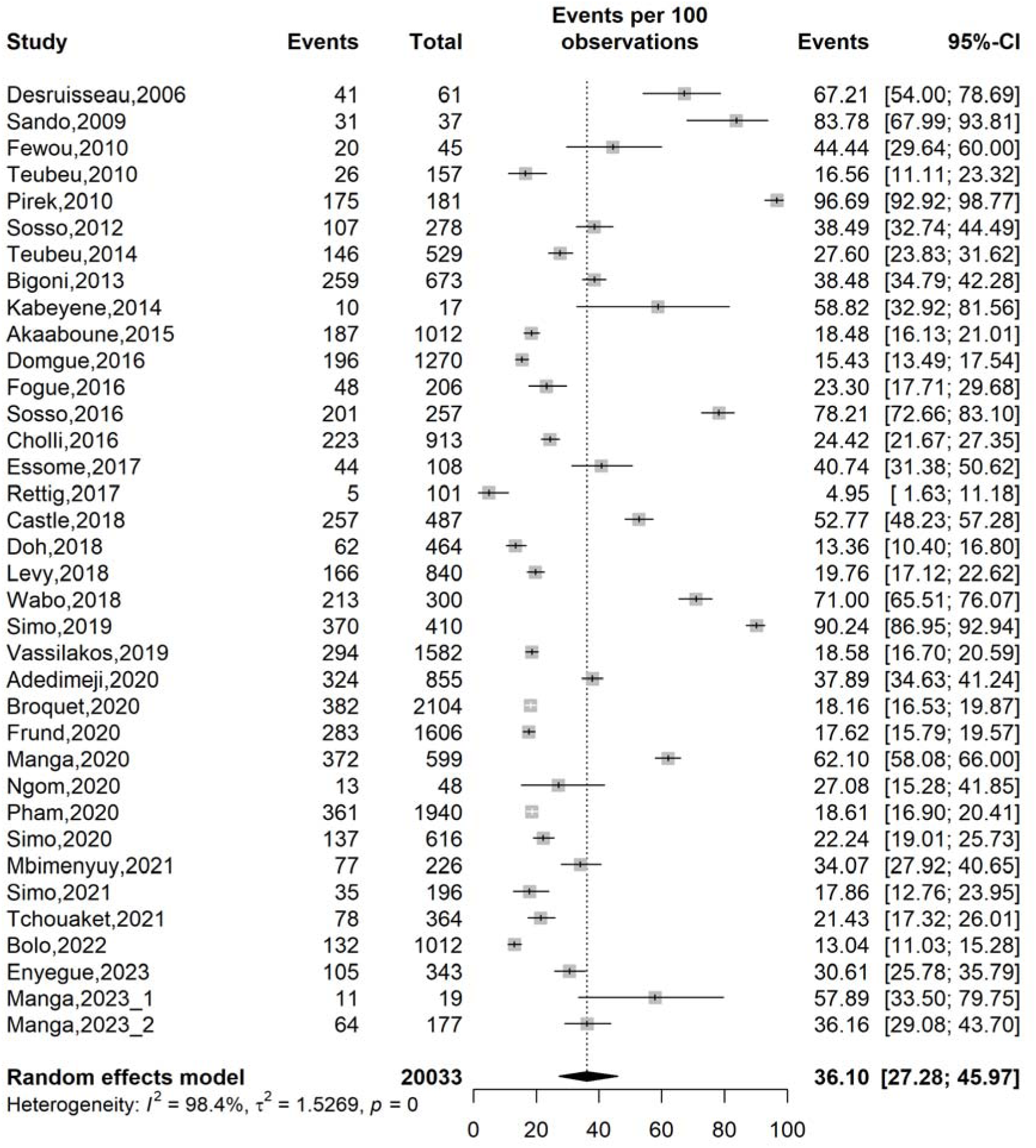
Pooled prevalence of human papillomavirus infection in Cameroon

The subgroup analysis reveals significant variation in prevalence based on study period, setting, region, and participant characteristics. The analysis identified a clear downward trend in HPV prevalence over time, which aligns with public health interventions. Studies conducted before 2014 (pre-vaccination era) showed the highest pooled prevalence at 55.65% (95% CI: 30.70–78.04%; 8 studies; n = 1,961). The period of 2014–2020, corresponding to HPV vaccination demonstration projects, saw a marked decrease to 32.21% (95% CI: 22.26–44.32%; 21 studies; n = 15,735). The most recent period (2021–2023: post-vaccine introduction) showed the lowest pooled prevalence at 26.98% (95% CI: 19.11–36.62%; 7 studies; n = 2,337). While this trend is promising, the subgroup difference was not statistically significant (*p* = 0.105) (**Table 2 and Supplementary Fig. 1**).

**Table 2.** Descriptive characteristics of included studies.

| Author | Year<sup>1</sup> | Design | Region | Setting | Sampling method | Type of participant | Age (years) | Median IQR or Mean SD | HPV diagnostic tool | Risk of bias | Sample size | Prevalence (%) |
| --- | --- | --- | --- | --- | --- | --- | --- | --- | --- | --- | --- | --- |
| Desruisseau,2006 [28] | 2006 | Cross-sectional | Centre and South-West | Hospital | Non-probalistic | Non-pregnant women | 18+ | NR | PCR-based reverse-line strip test for multiple HPV types | Low | 61 | 67.21 |
| Sando,2009 [29] | 2009 | Cross-sectional | Centre | Hospital | Non-probalistic | Women with pre/cancerous genital lesion | 28-55 | NR | PCR-based reverse-line strip test for multiple HPV types | Low | 37 | 83.78 |
| Fewou,2010 [30] | 2010 | Cross-sectional | Centre and Littoral | Hospital | Non-probalistic | Women with pre/cancerous genital lesion | 64-76 | 69 SD NR | Type specific PCR | Moderate | 45 | 44.44 |
| Teubeu,2010 [23] | 2010 | Cross-sectional | West | Hospital | Non-probalistic | Women | 30-75 | NR | Abbott RealTime High-Risk HPV assay | Low | 157 | 16.56 |
| Pirek,2010 [31] | 2010 | Cross-sectional | Centre and Littoral | Hospital | Non-probalistic | Women with pre/cancerous genital lesion | 29-87 | 52 IQR | Type specific PCR | Low | 181 | 96,69 |
| Sosso,2012 [32] | 2012 | Cross-sectional | Centre | Hospital | Non-probalistic | Sexually active non-pregnant women | 20-75 | 37.0 SD 3.0 | Abbott RealTime High-Risk HPV assay | Low | 278 | 38.49 |
| Teubeu,2014 [33] | 2012 | Cross-sectional | Centre and South-West | Community | Non-probalistic | Women | 30-65 | 41 IQR 36-50 | Abbott RealTime High-Risk HPV assay | Low | 529 | 27.60 |
| Bigoni,2013 [34] | 2013 | RCT | Centre | Community | Non-probalistic | Non-pregnant women | 25-65 | 41.4 SD 10.1 | Roche Cobas 4800 System | Low | 673 | 38.48 |
| Kabeyene,2014 [35] | 2014 | Cross-sectional | Centre | Hospital | Non-probalistic | Sexually active WLWH | 19-79 | 38.5 SD 9.9 | Abbott RealTime High-Risk HPV assay | Moderate | 17 | 58.82 |
| Akaaboune,2015 [36] | 2015 | Cross-sectional | West | Hospital | Non-probalistic | Non-pregnant women | 30-59 | 38.7 SD 5.6 | Cepheid GeneXpert | Low | 1,012 | 18.48 |
| Domgue,2016 [37] | 2016 | Cross-sectional | North-West | Community | Non-probalistic | Non-pregnant women | 30-65 | 44.7 SD 9.8 | careHPV | Low | 1,270 | 15.43 |
| Fogue,2016 [38] | 2016 | Cross-sectional | West | Hospital | Non-probalistic | Sexually active women | 16-50 | 28.1 SD 8.0 | Type specific PCR | Low | 206 | 23.30 |
| Sosso,2016 [39] | 2016 | Cross-sectional | Centre | Hospital | Non-probalistic | Sexually active non-pregnant women | 18+ | 37.0 SD 6.5 | Abbott RealTime High-Risk HPV assay | Low | 257 | 78.21 |
| Cholli,2016 [40] | 2016 | Cross-sectional | Littoral and North-West | Hospital and community | Non-probalistic | Non-pregnant women | 30-80 | 41.8 SD 9.9 | careHPV | Low | 913 | 24,42 |
| Essome,2017 [41] | 2017 | Cross-sectional | Littoral | Hospital | Non-probalistic | Women | 21-71 | 40.1 SD 10.0 | Indirect immunoperoxidase technique (P16INK4a) | Low | 108 | 40.74 |
| Rettig,2017 [42] | 2017 | Cross-sectional | North-West | Hospital | Non-probalistic | PLHIV | 24-69 | 42 IQR 37-48 | OraRisk (PCR-based) | Low | 101 | 4.95 |
| Castle,2018 [43] | 2018 | Cross-sectional | South-West | Hospital | Non-probalistic | WLWH | 25-59 | 42 IQR NR | Cepheid GeneXpert | Low | 487 | 52,77 |
| Doh,2018 [22] | 2018 | Cross-sectional | Centre | Hospital | Non-probalistic | Pregnant women | 25+ | NR | Roche Linear Array | Low | 464 | 13.36 |
| Levy,2018<br>[44] | 2018 | Cross-sectional | West | Community | Non-probalistic | Non-pregnant women | 30-49 | 39.4<br>SD 5.9 | Cepheid GeneXpert | Low | 840 | 19.76 |
| Wabo,2018<br>[45] | 2018 | Case-control | South-West | Hospital | Probabilistic | Women | 25-65 | 41.1<br>SD 10.5 | HPV High Risk Typing Real-TM | Low | 300 | 71.00 |
| Simo,2019<br>[46] | 2019 | Cross-sectional | Centre | Hospital | Non-probalistic | Women with pre/cancerous genital lesion | 20-84 | NR | Type spefic PCR | Low | 410 | 90.24 |
| Vassilakos,2019<br>[47] | 2019 | Cross-sectional | West | Hospital | Probabilistic | Non-pregnant women | 30-50 | 40<br>IQR 35-45 | Abbott RealTime High-Risk HPV assay | Low | 1,582 | 18.58 |
| Adedimeji,2020<br>[48] | 2020 | Cross-sectional | South-West | Hospital | Non-probalistic | Women | 19-59 | NR | Cepheid GeneXpert | Low | 855 | 37.89 |
| Broquet,2020<br>[49] | 2020 | Cross-sectional | West | Community | Non-probalistic | Non-pregnant women | 30-49 | 40<br>IQR 35-45 | Cepheid GeneXpert | Low | 2,104 | 18.16 |
| Frund,2020<br>[50] | 2020 | Cross-sectional | West | Community | Non-probalistic | Women | NR | 38.3<br>IQR 27-46 | Cepheid GeneXpert | Low | 1,606 | 17.62 |
| Manga,2020<br>[21] | 2020 | Cross-sectional | West, Centre and Littoral | Hospital | Non-probalistic | Female sex worker | 30+ | NR | AmpFire Assay | Low | 599 | 62.10 |
| Ngom,2020<br>[51] | 2020 | Cross-sectional | Centre and Littoral | Hospital | Non-probalistic | Patient with head and neck cancers | 20-78 | 49.0<br>SD NR | Abbott RealTime High-Risk HPV Assay | Low | 48 | 27.08 |
| Pham,2020<br>[52] | 2020 | Cross-sectional | West | Community | Non-probalistic | Non-pregnant women | 30-49 | 40.2<br>SD 5.9 | Cepheid GeneXpert | Low | 1,940 | 18.61 |
| Simo,2020<br>[53] | 2020 | Cross-sectional | Centre | Hospital | Non-probalistic | Women | 20-65 | NR | Type specific PCR | Low | 616 | 22.24 |
| Mbimenyuy,2021<br>[54] | 2021 | Cross-sectional | Centre | Hospital | Non-probalistic | Sexually active non-pregnant women | 19+ | 44.0<br>SD 10.8 | Roche Linear Array | Low | 226 | 34.07 |
| Simo,2021<br>[55] | 2021 | Cross-sectional | Littoral | Hospital | Non-probalistic | Sexually active women | 20-70 | NR | Abbott RealTime High-Risk HPV assay | Low | 196 | 17.86 |
| Tchouaket,2021<br>[56] | 2021 | Cross-sectional | Centre and Littoral | Hospital | Non-probalistic | Sexually active non-pregnant women | 18+ | 41<br>IQR 34-50 | Abbott RealTime High-Risk HPV assay | Low | 364 | 21.43 |
| Bolo,2022<br>[57] | 2022 | Cohort | West | Community | Non-probalistic | Non-pregnant women | 30-49 | 37.4<br>SD 3.9 | Roche Cobas 4800 System | Low | 1,012 | 13.04 |
| Enyegue,2023<br>[19] | 2023 | Cross-sectional | Centre | Hospital | Non-probalistic | Sexually active women | 15-65 | NR | QIAGEN Multiplex PCR Kit | Low | 343 | 30.61 |
| Manga,2023_1<br>[18] | 2023 | Cross-sectional | Littoral | Community | Non-probalistic | WSW | 19-40 | 29.2<br>SD 5.3 | AmpFire Assay | Low | 19 | 57.89 |
| Manga,2023_2<br>[17] | 2023 | Cross-sectional | North-West | Hospital | Non-probalistic | WLWH | 25+ | NR | AmpFire Assay | Low | 177 | 36.16 |

1: Year of study completion; RCT: Randomized control trial; HPV: Human papillomavirus; WLWH: Women living with HIV; PLVIH; People living with HIV; NR: Not reported; IQR: Interquartile range; SD: Standard deviation; WSW: Women having sex with women; PCR: Polymerase chain reaction;

The study setting proved to be a source of heterogeneity of the reported prevalence, with a statistically significant difference (*p* = 0.009). Hospital-based studies, reported nearly double the prevalence (41.94%, 95% CI: 29.99–49.78%, 26 studies, n = 9,127) compared to community-based screenings (22.08%, 95% CI: 16.62–28.72%, 9 studies, n = 9,993). No significant differences were found based on study design (*p* = 0.903) or sampling method (*p* = 0.646) (**Table 2 and Supplementary Fig. 3**).

The geographic analysis revealed a statistically significant regional disparity (*p* < 0.001). The highest prevalence was found in the South-West region (54.08%, 95% CI: 38.24–69.14%, 3 studies, n=1,642) and the Centre region (49.47%, 95% CI: 30.69–68.41%, 10 studies, n=3,321). Conversely, the North-West (15.52%; 95% CI: 5.85-35.22; 3 studies; n = 1,548) and the West (17.95%, 95% CI: 16.58–19.41%, 9 studies, n = 10,459) regions had the lowest prevalence (**Table 2 and Supplementary Fig. 4 and 5**).

Important disparities were observed across different participant populations (*p*<0.001). The pooled prevalence among women with pre-cancerous genital lesions was high at 85.53% (95% CI: 61.72–95.59%, 4 studies, n = 673). Female sex workers also showed a very high prevalence of 62.10% (95% CI: 58.08–66.00%, 1 study, n = 599). PLHIV had a pooled prevalence of 31.92% (95% CI: 11.69–62.43%, 4 studies, n = 782). In contrast, women from the general population had a significantly lower pooled prevalence of 28.50% (95% CI: 22.40–35.50%, 26 studies, n = 17,931) (**Table 2 and Supplementary Fig. 7**).

### 3.4. Publication bias and sensitivity analysis

Asymmetric funnel plot suggested potential publication bias. In addition, the Egger’s (*p* = 0.018) and Begg’s (*p* = 0.026) test revealed significant risk of publication bias. We therefore performed the trim and fill analysis which imputed nine potentially unpublished studies. The adjusted pooled HPV infection prevalence revealed a non-significant declined from 36.10% (95% CI: 27.28-45.97; *I^2^* = 98.4%) to 23.62% (15.79-33.77; *I^2^* = 98.9%) (**Fig. 3 and 4**).

**Fig. 3.**
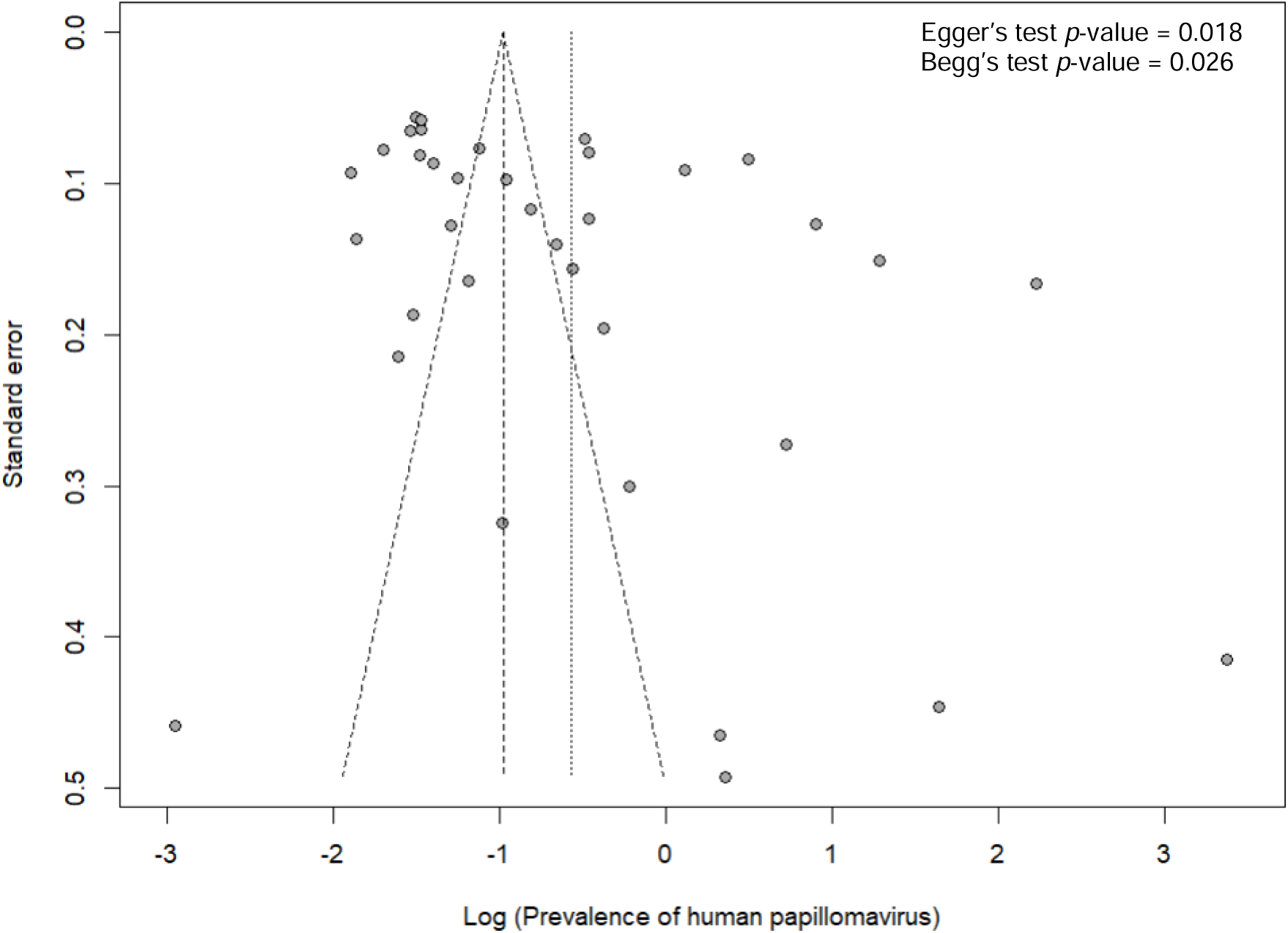
Funnel plot assessing the risk of publication bias of the pooled prevalence of human papillomavirus infection in Cameroon

**Fig. 4.**
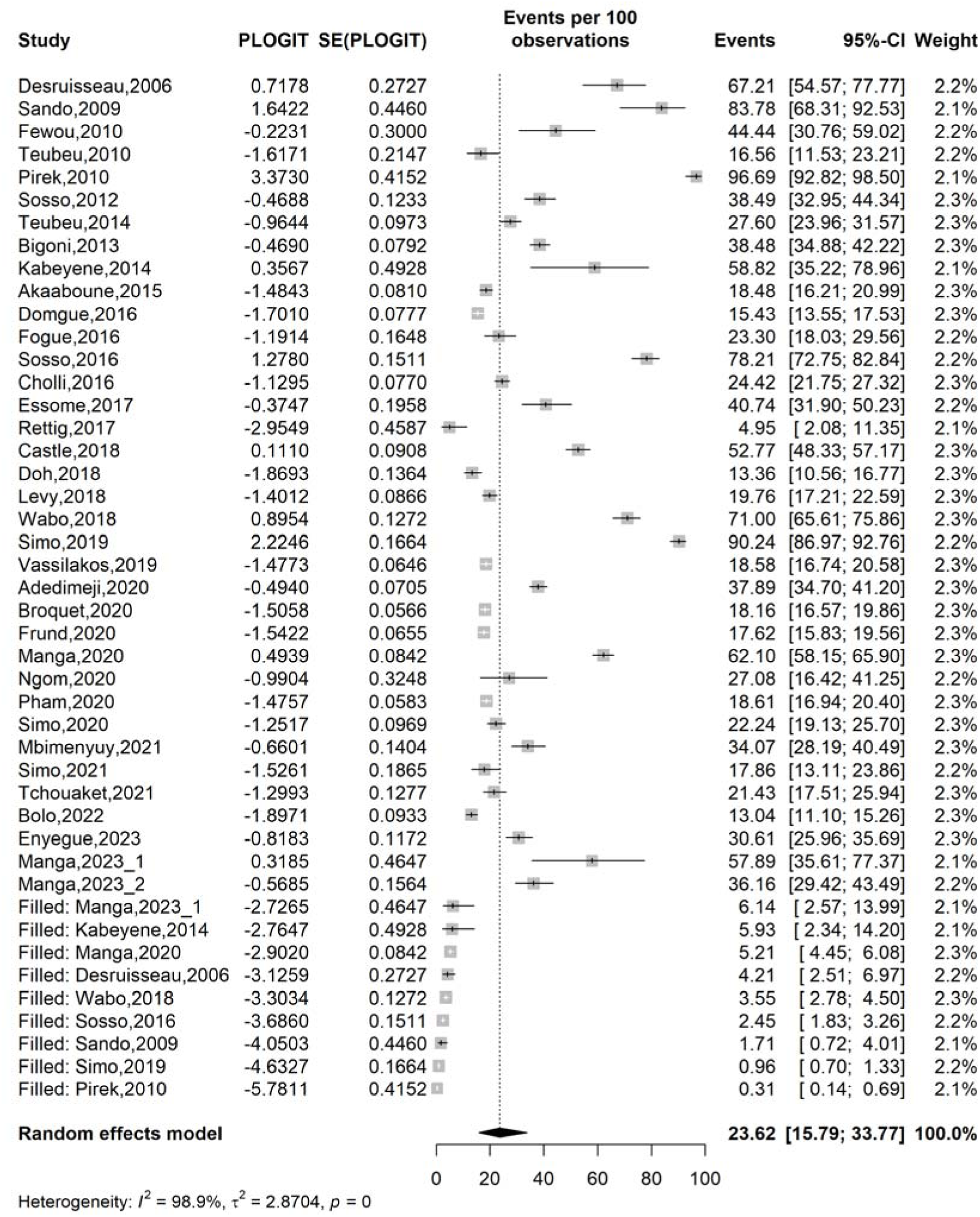
Trim and fill analysis of the pooled prevalence of human papillomavirus infection in Cameroon

The sensitivity analysis demonstrated no study significantly influenced the overall pooled HPV infection prevalence estimate (**Fig. 5**).

**Fig. 5.**
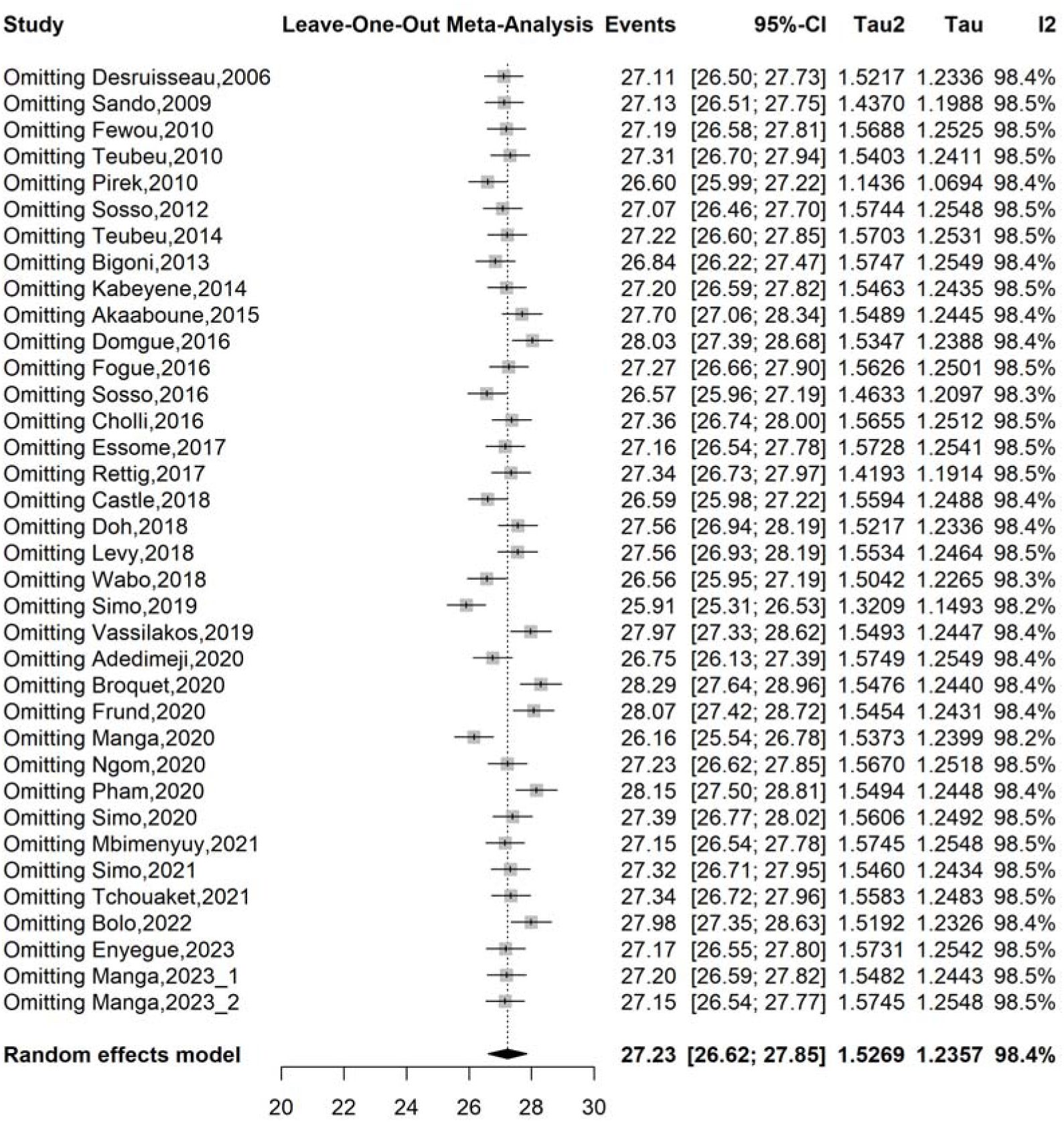
Sensitivity analysis of the pooled prevalence of human papillomavirus infection in Cameroon

### 3.5. Determinant of human papillomavirus infection

#### Age

Women aged less than 40 years were 31% significantly more at risk of HPV infection than their counterpart (OR = 1.31; 95% CI: 1.14-1.49; 7 reports) **(Table 3 and Supplementary Fig. 8).**

**Table 3.** Subgroup meta-analysis of the pooled prevalence of human papillomavirus infection in Cameroon.

| Subgroup | Sample size | Prevalence<sup>1</sup> (%) | 95% CI limits<sup>1</sup> |  | Number of studies | Heterogeneity statistic<sup>1</sup> |  | Subgroup difference |
| --- | --- | --- | --- | --- | --- | --- | --- | --- |
|  |  |  | Lower | Upper |  | I<sup>2</sup>(%) | p-value |  |
| **Study period (year)<sup>2</sup>** |  |  |  |  |  |  |  | 0.105 |
| └ 2014 | 1,961 | 55.65 | 30.70 | 78.04 | 8 | 96.1 | └ 0.001 |  |
| 2014-2020 | 15,735 | 32.21 | 22.26 | 44.32 | 21 | 98.9 | └ 0.001 |  |
| 2021-2023 | 2,337 | 26.98 | 19.11 | 36.62 | 7 | 94.6 | └ 0.001 |  |
| **Study design** |  |  |  |  |  |  |  | 0.903 |
| Cross-sectional | 18,048 | 35.94 | 26.94 | 46.33 | 33 | 98.3 | └ 0.001 |  |
| Other | 1,985 | 37.92 | 14.38 | 68.96 | 3 | 99.4 | └ 0.001 |  |
| **Study setting** |  |  |  |  |  |  |  | **0.009** |
| Community | 9,993 | 22.08 | 16.62 | 28.72 | 9 | 96.4 | └ 0.001 |  |
| Hospital | 9,127 | 41.94 | 29.99 | 49.78 | 26 | 98.4 | └ 0.001 |  |
| Hospital & community | 913 | 24.52 | 21.67 | 27.35 | 1 | - | - |  |
| **Region** |  |  |  |  |  |  |  | □ **0.001** |
| Centre | 3,321 | 49.47 | 30.69 | 68.41 | 10 | 98.5 | └ 0.001 |  |
| Littoral | 323 | 35.14 | 18.73 | 56.00 | 3 | 92.1 | └ 0.001 |  |
| North-West | 1,548 | 15.52 | 5.85 | 35.22 | 3 | 96.2 | └ 0.001 |  |
| South-West | 1,642 | 54.08 | 38.24 | 69.14 | 3 | 98.0 | └ 0.001 |  |
| West | 10,459 | 17.95 | 16.58 | 19.41 | 9 | 66.2 | 0.003 |  |
| Centre & Littoral | 638 | 55.01 | 16.24 | 88.52 | 4 | 97.5 | └ 0.001 |  |
| Centre & South-West | 590 | 46.08 | 20.89 | 73.45 | 2 | 97.0 | └ 0.001 |  |
| Centre, Littoral, & West | 599 | 62.10 | 58.08 | 66.00 | 1 | - | - |  |
| Littoral & North-West | 913 | 24.52 | 21.67 | 27.35 | 1 | - | - |  |
| Multi-region<sup>3</sup> | 2,740 | 49.43 | 26.46 | 72.65 | 8 | 98.1 | └ 0.001 |  |
| **Sampling method** |  |  |  |  |  |  |  | 0.646 |
| Non-probabilistic | 18,151 | 35.72 | 26.71 | 45.87 | 34 | 98.3 | └ 0.001 |  |
| Probabilistic | 1,882 | 42.68 | 12.54 | 79.45 | 2 | 99.6 | └ 0.001 |  |
| **Participant** |  |  |  |  |  |  |  | □ **0.001** |
| Women from general population | 17,931 | 28.50 | 22.40 | 35.50 | 26 | 97.8 | └ 0.001 |  |
| PLHIV | 782 | 31.92 | 11.69 | 62.43 | 4 | 94.4 | └ 0.001 |  |
| Women with pre/cancerous genital lesions | 673 | 85.53 | 61.72 | 95.59 | 4 | 95.5 | └ 0.001 |  |
| Patient with head & neck cancers | 48 | 27.08 | 15.28 | 41.85 | 1 | - | - |  |
| Female sex worker | 599 | 62.10 | 58.08 | 66.00 | 1 | - | - |  |

<sup>1</sup> Random effects model; <sup>2</sup> └ 2014 = Pre-HPV vaccination in Cameroon, 2014-2020 = HPV vaccination demonstration in Cameroon, 2021-2023 = post-HPV vaccine introduction in Cameroon; <sup>3</sup> Multi-region included studies conducted in more than one of following regions (Centre, Littoral, South-West, North-West, and West); CI: Confidence interval; PLHIV: People living with human immunodeficiency virus (HIV)

#### Marital status

Unmarried women were 43% significantly more likely to be HPV positive than those living in partnership (OR = 1.43; 95% CI: 1.24-1.64; 15 reports) (**Table 3 and Supplementary Fig. 10).**

#### Number of sexual partners

Women who reported having five or more sexual partners were 26% significantly more like to get infected with HPV than those with less than five partners (OR = 1.26; 95% CI: 1.05-1.51; 2 reports) (**Table 3 and Supplementary Fig. 15-16**).

#### Contraception use

Women who reported using hormonal contraceptives or condom were 39% (OR = 1.61; 95% CI: 1.19-2.17) and 61% (OR = 1.21; 95% CI: 1.09-1.52) significantly more at risk of HPV infection than those who reported no contraception respectively (**Table 3 and Supplementary Fig. 17-18**).

#### Parity

Women who reported less than 4 normal deliveries were more likely to be diagnosed with HPV than those who reported higher deliveries (OR = 1.29; 95% CI: 1.09-1.52) (**Table 3 and Supplementary Fig 19**).

#### HIV status

PLHIV were 92% more likely to be HPV positive than women HIV negative (OR = 1.92; 1.24-2.98) (**Table 3 and Supplementary Fig. 20**).

#### CD4 count

The risk of being HPV positive was twice higher among individuals with less 500 CD4 cell/mm^3^ than their counterpart (OR = 2.00; 1.02-3.95) (**Table 3 and Supplementary Fig. 21**).

#### HIV viral load

PLHIV with high viral load (1000 + copies/mL) were 2.12 times more likely to get infected with HPV than those with lower viral load (OR = 2.12; 1.27-3.53) (**Table 3 and Supplementary Fig. 22**).

## 4. Discussion

This systematic review and meta-analysis provide the most comprehensive synthesis to date of the prevalence and determinants of HPV infection in Cameroon. Based on 36 studies including 20,033 participants, the pooled HPV prevalence was high (36.1%), highlighting HPV as a major public health concern in the country. However, substantial heterogeneity was observed across studies, reflecting differences in study populations, geographic regions, settings, diagnostic methods, and time periods. Our findings were comparable to estimates from other sub-Saharan African settings, but remained markedly higher than those reported in high-income countries [11,58,59]. This disparity likely reflects differences in sexual and reproductive health profiles, limited access to organized cervical cancer screening, and the continued high prevalence of HIV infection in the African region [60,61].

In Cameroon, cervical cancer screening is predominantly opportunistic, with many women being screened for the first time only after the onset of symptoms. This pattern likely contributes to the higher HPV prevalence reported, especially in hospital-based studies. In addition, while the HPV vaccine was introduced in Cameroon around 10 years ago, uptake has remained limited [62].

Further in our analysis, we observed a gradual decline in HPV prevalence over time, with the highest estimates reported in studies conducted before 2014 and the lowest in most recent studies. Although the differences across study periods were not statistically significant, this downward trend is epidemiologically significant. It may reflect the impact of public health interventions such as HPV vaccination projects, increased cervical cancer awareness, and the availability of HPV-based screening, which likely influenced the adoption of lower-risk sexual practices in Cameroon [63–66]. Nevertheless, the persistence of a relatively high prevalence in the post-vaccine introduction period suggests that vaccination coverage remains insufficient to substantially reduce population-level transmission, emphasizing the need to strengthen vaccine implementation [67].

Also, the study setting emerged as an important source of variation in HPV prevalence. Hospital-based studies consistently reported a higher prevalence than community-based studies, likely reflecting the selection of women with symptoms, abnormal screening results, or underlying conditions that increase susceptibility to HPV infection. Similar patterns have been reported elsewhere in sub-Saharan Africa and highlight the influence of referral and selection bias in facility-based studies [68,69]. Community-based studies may better approximate the true population burden of HPV and reinforce the importance of expanding outreach screening programs.

HPV prevalence varies across Cameroon regions even though with high heterogeneity. These differences may be due to factors like urbanization, healthcare access, HIV rates, sexual behavior, and where diagnostic and research facilities are located [70–72]. Also, areas with many reference hospitals like Centre region may be overrepresented in surveillance activities, potentially leading to an overestimation of HPV prevalence in these regions. However, the highest prevalence observed in the South-West region may reflect socio-cultural component that may influence sexual behavior across communities in this specific region. Evidences from this region describe a high prevalence of risky sexual behaviors among both adolescents and adults and some reasons reported included poverty, lack of sexual education, disbelief in the existence of sexually transmitted infectious disease like HIV/AIDS, neglect of voluntary counseling and testing services [73–75]. Our findings highlight the importance of developing region-specific strategies to reinforce HPV prevention in Cameroon.

HPV infection rates were much higher among groups at greater risk. Women with pre-cancerous or cancerous genital lesions had very high rates of HPV, which supports the known link between long-term HPV infection and cervical cancer development [76,77]. Female sex workers also had higher rates, likely due to more frequent exposure from multiple sexual partners [78]. People living with HIV were more often infected with HPV, which matches evidence that HIV-related immune suppression makes it harder to clear HPV and increases the risk of persistent infection and serious lesions [79–81]. The strong links between HPV infection, low CD4 counts, and high HIV viral load show how important immune health is for controlling HPV and preventing disease progression [82].

This meta-analysis found that HPV infection is linked to several social, behavioral, reproductive, and clinical factors. Younger women, especially those under 40, had 31% higher odds of HPV infection than older women, which is consistent with the natural history of HPV acquisition, as infections are most common shortly after sexual debut and during periods of higher sexual activity [83]. Unmarried women were 43% more likely to be HPV positive than those in stable partnerships, possibly because of more partner changes. Sexual behavior was important, as women with five or more lifetime sexual partners had a higher chance of HPV infection than those with fewer partners, which matches known HPV transmission patterns [83]. The link between contraceptive use and HPV infection should be viewed carefully. Women who used hormonal contraceptives or condoms had higher odds of HPV infection, likely reflecting sexual behavior rather than a direct effect of contraception [84]. This association may be explained by the fact that contraception reduces the fear of unintended pregnancy; this may lead to a higher number of sexual partners, thereby increasing the odds of HPV transmission.

Reproductive history also played a role; women with fewer than four deliveries were more likely to test positive for HPV than those with more children, possibly due to differences in age or sexual behavior. Women with more children might have a more responsible sexual behavior than their counterpart [85]. HIV infection was also a strong predictor of HPV, as people living with HIV were almost twice as likely to have HPV compared to HIV-negative women. Among PLHIV, immune status played a critical role as individuals with CD4 cell counts below 500 cells/mm³ had a twofold higher odd of HPV infection, and those with high HIV viral loads (≥1000 copies/mL) were also more likely to be HPV positive, underscoring the importance of immune suppression and poor viral control in HPV persistence. Our results are consistent with previous research that showed HIV infection favors the development of high-risk HPV genotype [82].

## 5. Limitations

This review has several important limitations. The primary studies interested five regions over ten and might not totally reflect the situation in the whole country. There was a high variation between studies because of differences in populations, settings, regions, and diagnostic methods, which could affect how precise the pooled estimates were. Most studies were cross-sectional, hospital-based, and used non-probability sampling, which might lead to an overestimate of HPV prevalence and make it harder to apply the findings more broadly. We found evidence of publication bias. While the trim-and-fill adjustment lowered the pooled prevalence, this method is based on assumptions and may not fully address unpublished data. Also, some subgroup and determinant analyses relied on only a few studies, and behavioral variables were self-reported. Because of these factors, the results should be interpreted with caution.

## 6. Conclusions

HPV infection remains a major public health concern in Cameroon. While the pooled prevalence was 36.1%, adjustment for publication bias using the trim-and-fill method yielded a lower estimate of 23.6%. Our results show that Cameroon urgently needs stronger HPV prevention efforts to meet the “90-70-90” targets by 2030. Expanding HPV vaccination, adding HPV testing to regular cervical cancer screening, and focusing on high-risk groups like young women, female sex workers, and people living with HIV are key steps to lowering morbidity and mortality from HPV complications. Moreover, prevention strategies that educate population and raise awareness about the disease including its oncogenic potential will help move toward cervical cancer elimination objectives in Cameroon. Future studies should determine HPV prevalence in the five unexplored regions including Far-North, North, Adamawa, East and South regions of Cameroon.

## Data Availability

All data generated or analyzed during this study are included in this published article and supplemental material.

## 7. Abbreviations

CI Confidence interval

HIV Human immunodeficiency virus

HPV Human papillomavirus

HR-HPV High-risk human papillomavirus

IQR Interquartile range

MeSH Medical subject headings

NR Not reported

OR Odds ratio

PCR Polymerase chain reaction

PLHIV people living with HIV

PRISMA Preferred reporting items for systematic reviews and meta-analysis

RCT Randomized control trial

SD Standard deviation

SSA Sub-Saharan Africa

WLWH Women living with HIV

WSW Women having sex with women

## 8. Declarations

### Ethical approval and consent to participate

Not applicable.

### Consent for publication

Not applicable.

### Availability of data and materials

The sources of data supporting this systematic review are available in the reference. All data generated or analyzed during this study are included in this published article and supplemental material.

### Competing interests

All authors declare no conflicts of interest and have approved the final version of the article.

### Code availability

All codes used in this study are available upon request from the corresponding author.

### Funding source

This research did not receive any specific grant from funding agencies in the public, commercial or not-for-profit sectors.

### Author contributions

FZLC conceived the original idea of the study; FZLC and RT conducted the literature search; FZLC, RT, ELMB and DGN selected the studies, extracted the relevant information, and synthesized the data; FZLC performed the analyses; FZLC, RT, CA, and SD wrote the first draft of the manuscript. All authors critically reviewed and revised successive drafts of the manuscript. All authors read and approved the final manuscript.

## Acknowledgements

None.

## Notes

### Competing Interest Statement

The authors have declared no competing interest.

### Author Declarations

The sources of data supporting this systematic review are available in the reference list.

