## Additional file for "HPV prevalence and associated factors in Cameroon: a systematic review and meta-analysis"

**Supplementary Table 1** Searching strategy by database

| **Database** | **Search string** | **Number of entries** |
| --- | --- | --- |
| **Pubmed** | ("human papillomavirus"[tiab] OR HPV[tiab] OR "cervical cancer"[tiab] OR "uterine cervical neoplasms"[Mesh]) AND (Cameroon[tiab] OR "Cameroon"[Mesh]) | 121 |
| **Scopus** | TITLE-ABS-KEY (("human papillomavirus" OR HPV OR "uterine cervical neoplasm") AND (Cameroon OR Cameroonian)) | 120 |
| **Web of sciences** | TS= ("human papillomavirus" OR HPV OR "cervical cancer" OR OR "uterine cervical cancer") AND TS= (Cameroon OR Cameroonian) | 217 |
| **Embase** | ('human papillomavirus':ti,ab,kw OR 'HPV':ti,ab,kw OR 'cervical cancer':ti,ab,kw OR 'uterine cervical neoplasms':ti,ab,kw) AND ('Cameroon':ti,ab,kw OR 'Cameroonian':ti,ab,kw) | 196 |
| **Cochrane Library** | human papillomavirus OR HPV OR cervical cancer) AND Cameroon OR Cameroun | 37 |
| **AJOL** | human papillomavirus OR HPV OR cervical cancer) AND Cameroon OR Cameroun | 33 |
| **Health Sciences and Disease** | (human papillomavirus OR HPV OR cervical cancer) AND (Cameroon OR Cameroun) | 9 |

**Subgroup analysis**

**Study period**

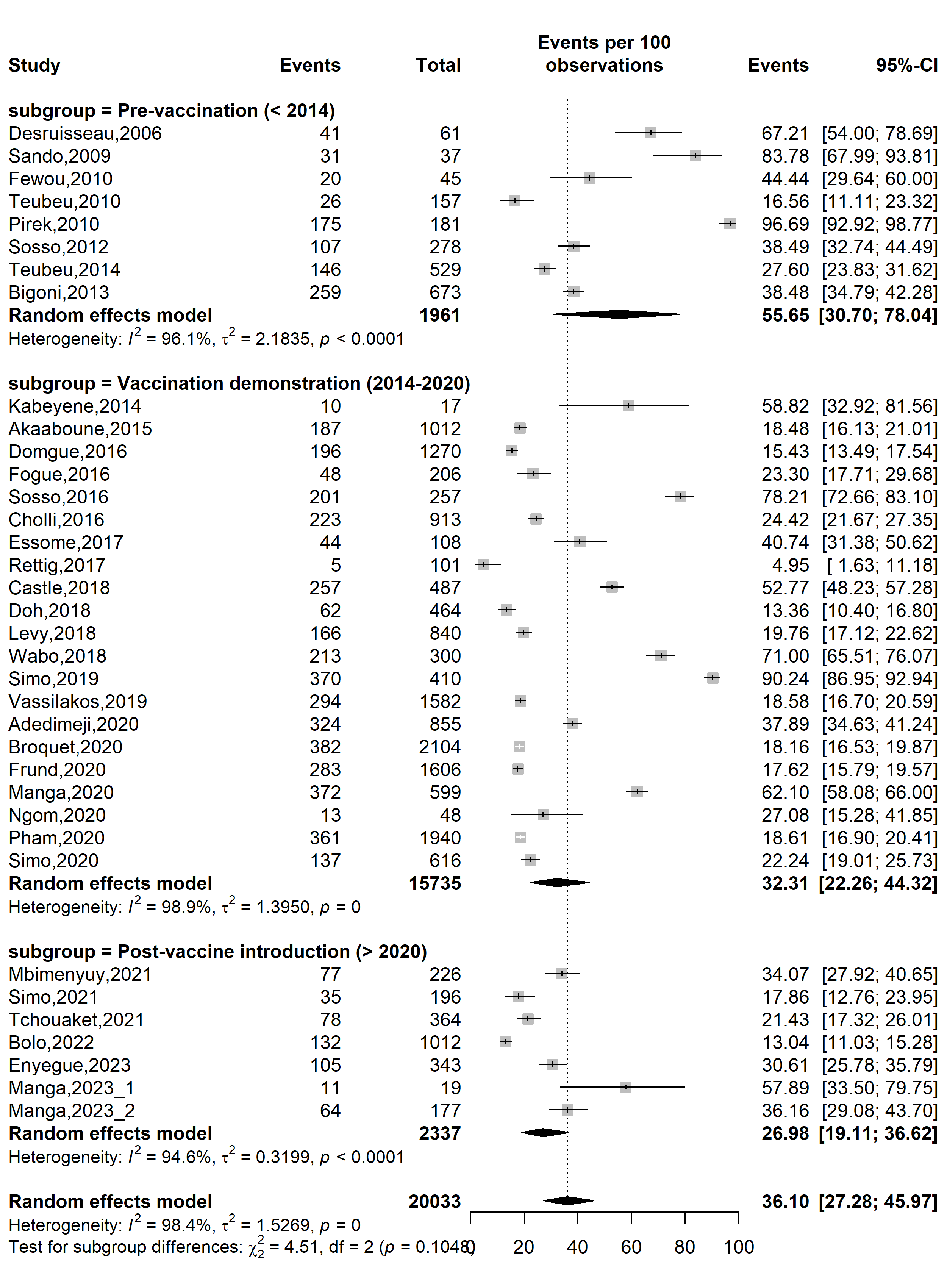

**Supplementary Fig. 1** Pooled HPV prevalence according to specific HPV vaccine introduction timeframe in Cameroon

**Study design**

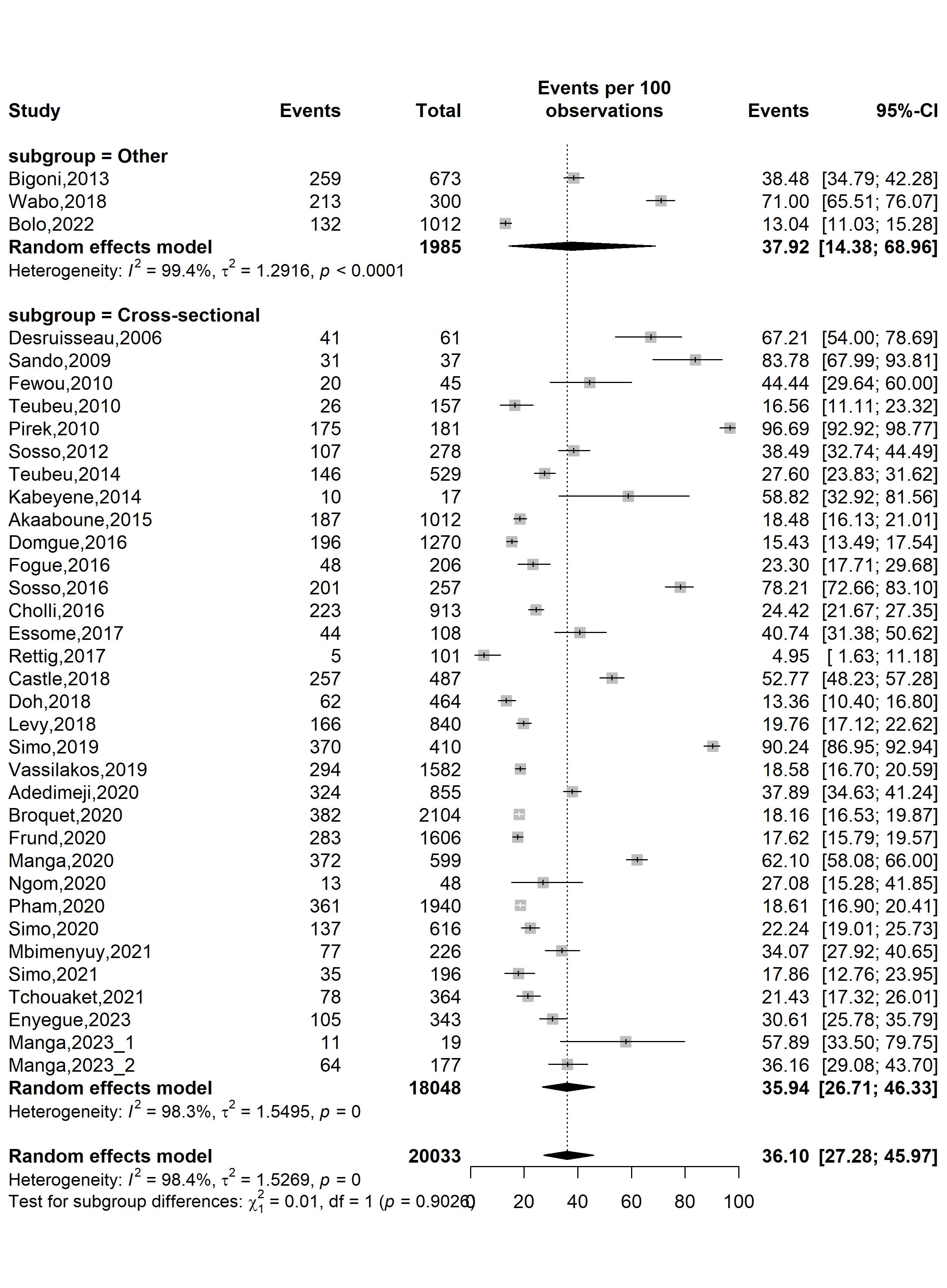

**Supplementary Fig. 2** Pooled HPV prevalence in Cameroon by study design

**Study setting**

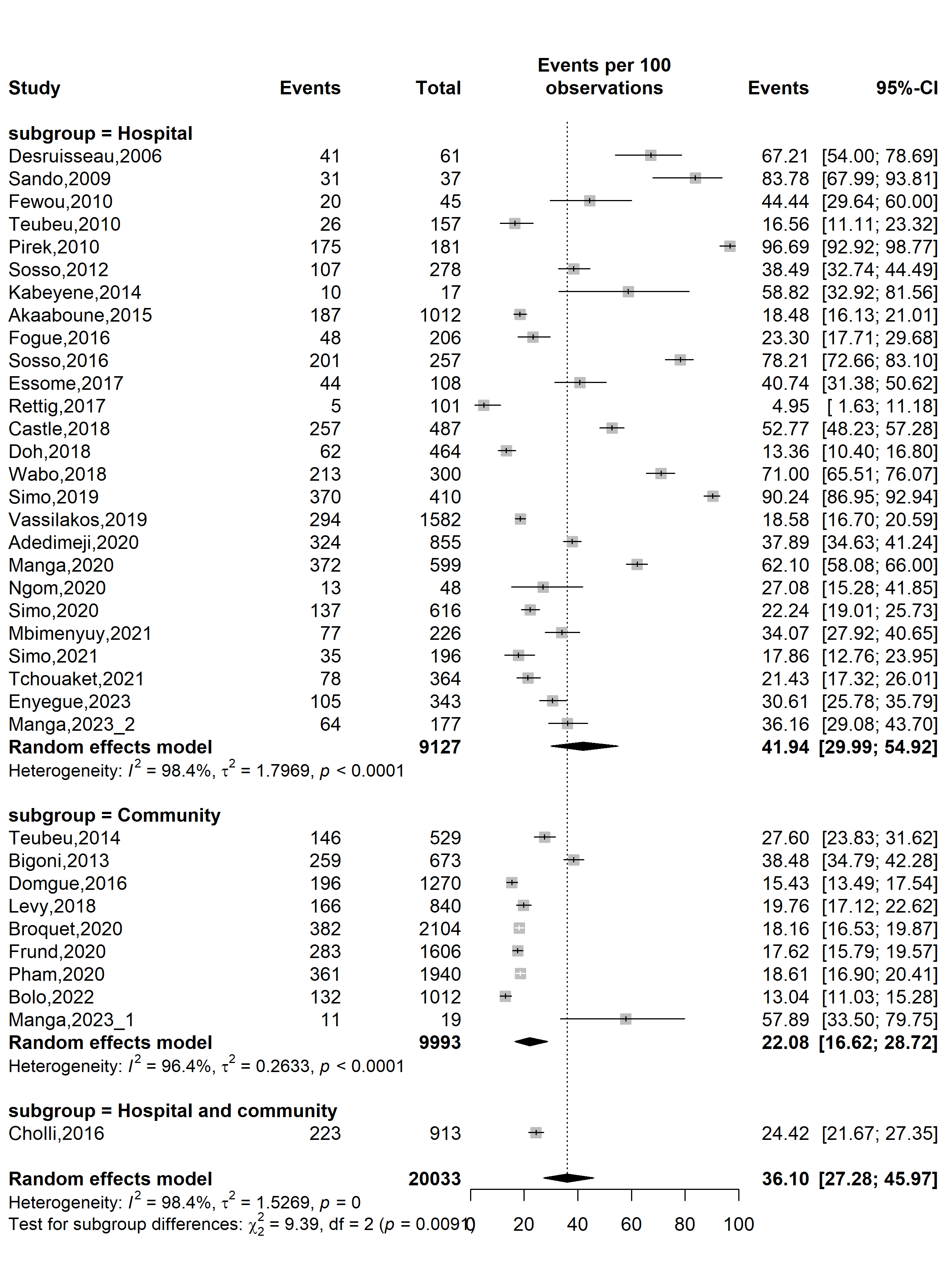

**Supplementary Fig. 3** Pooled HPV prevalence in Cameroon by study setting

**Study site**

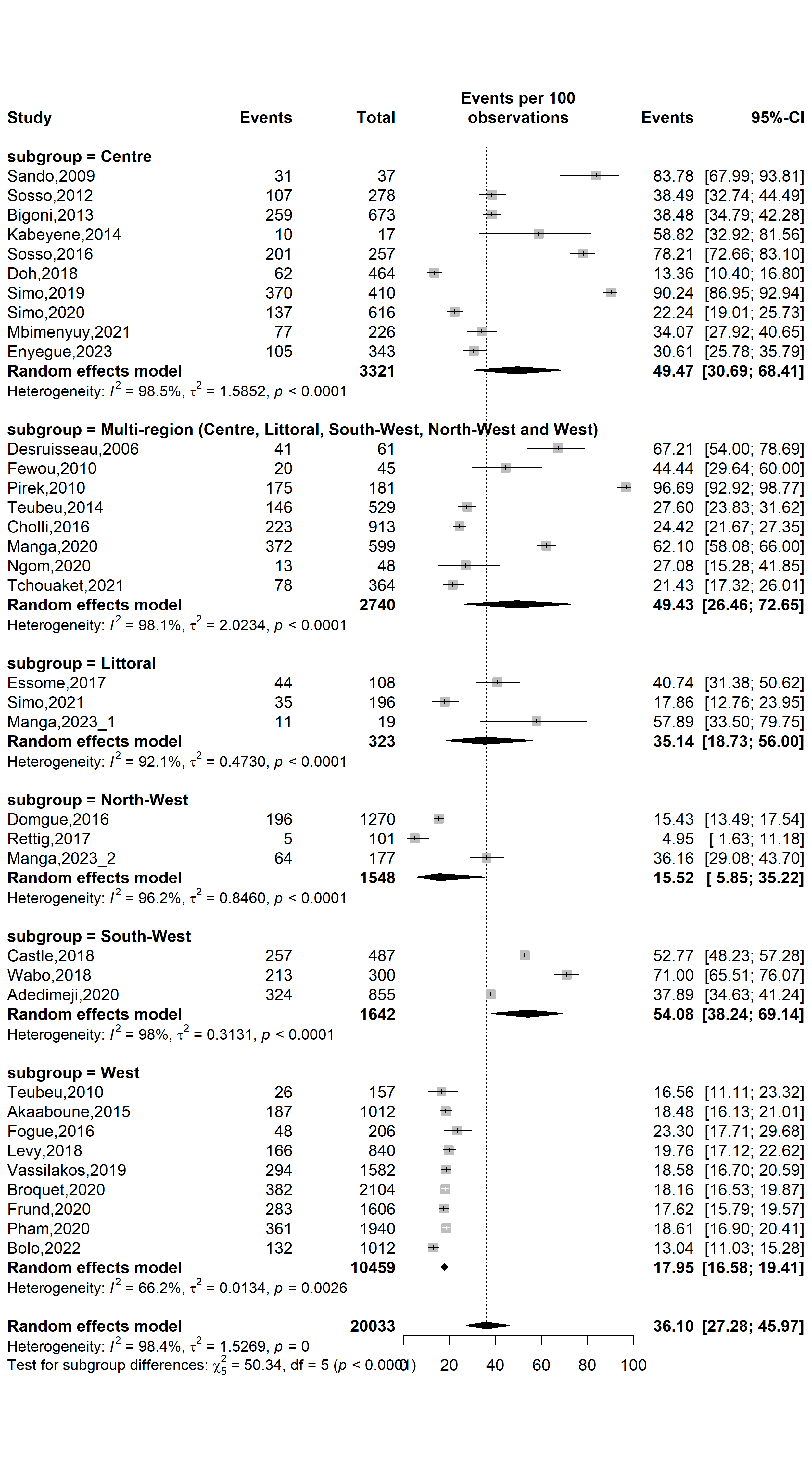

**Supplementary Fig. 4** Pooled HPV prevalence in Cameroon by study site 1

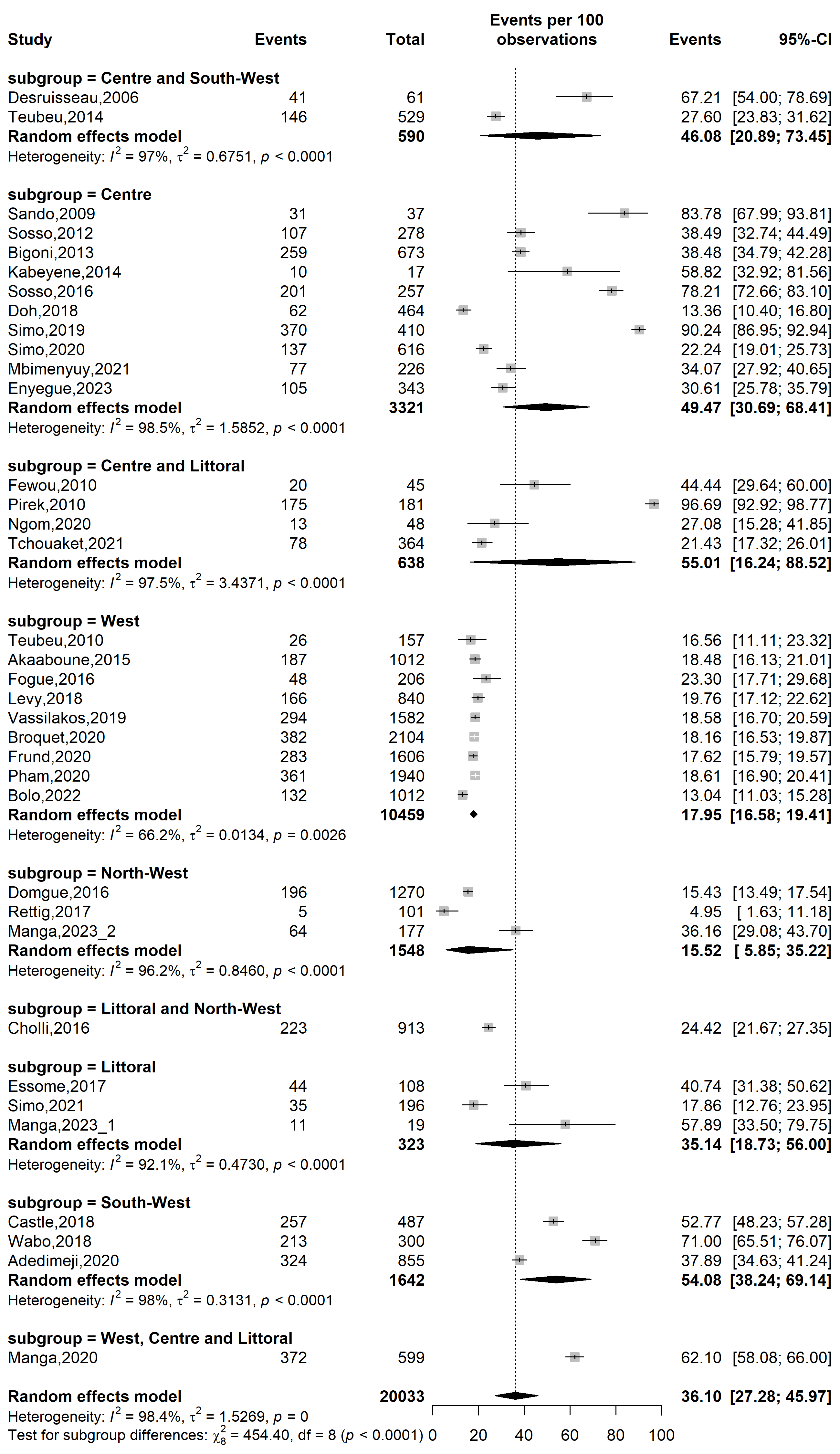

**Supplementary Fig. 5** Pooled HPV prevalence in Cameroon by study site 2

**Sampling method**

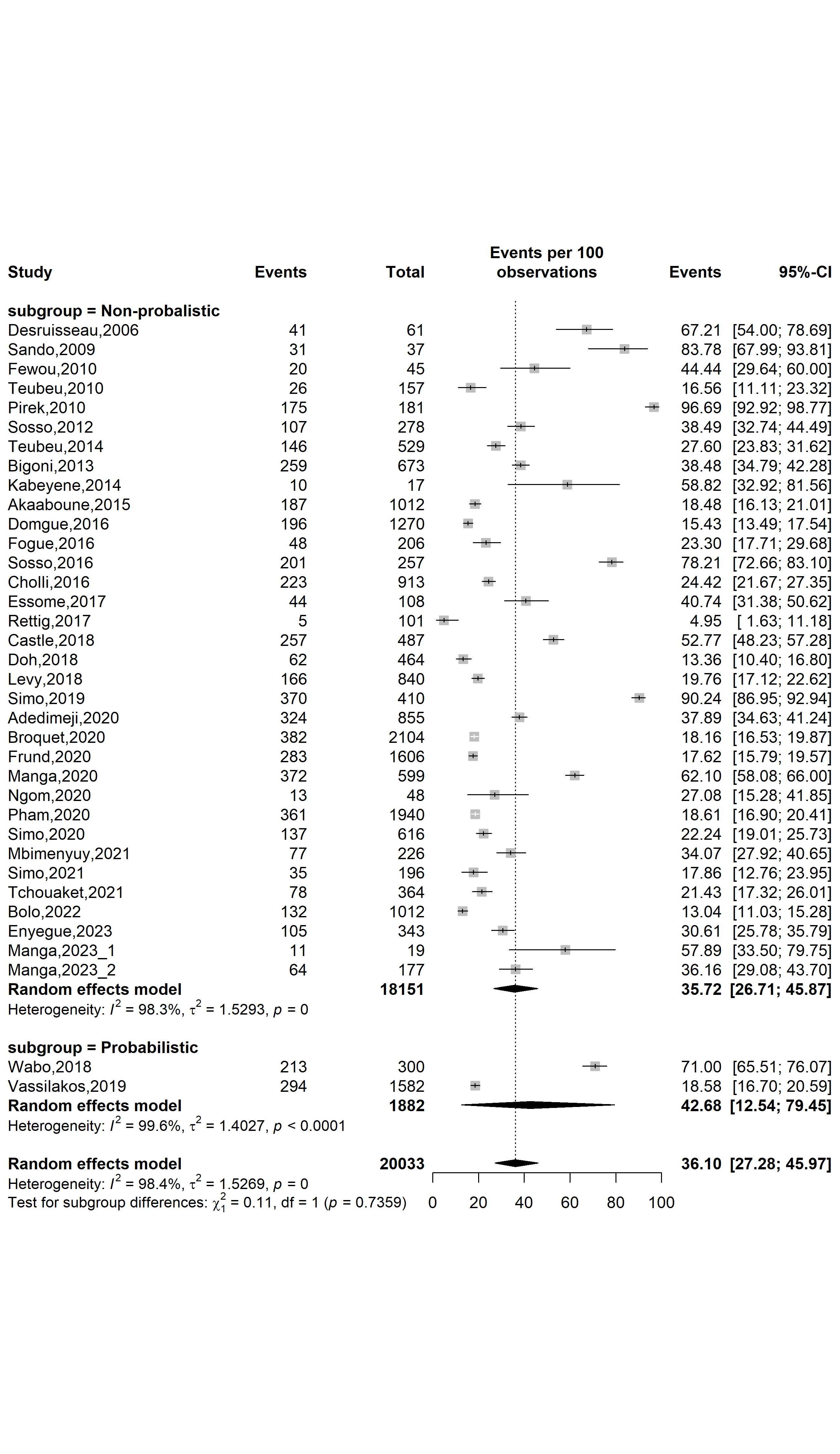

**Supplementary Fig. 6** Pooled HPV prevalence in Cameroon by sampling method

**Participant**

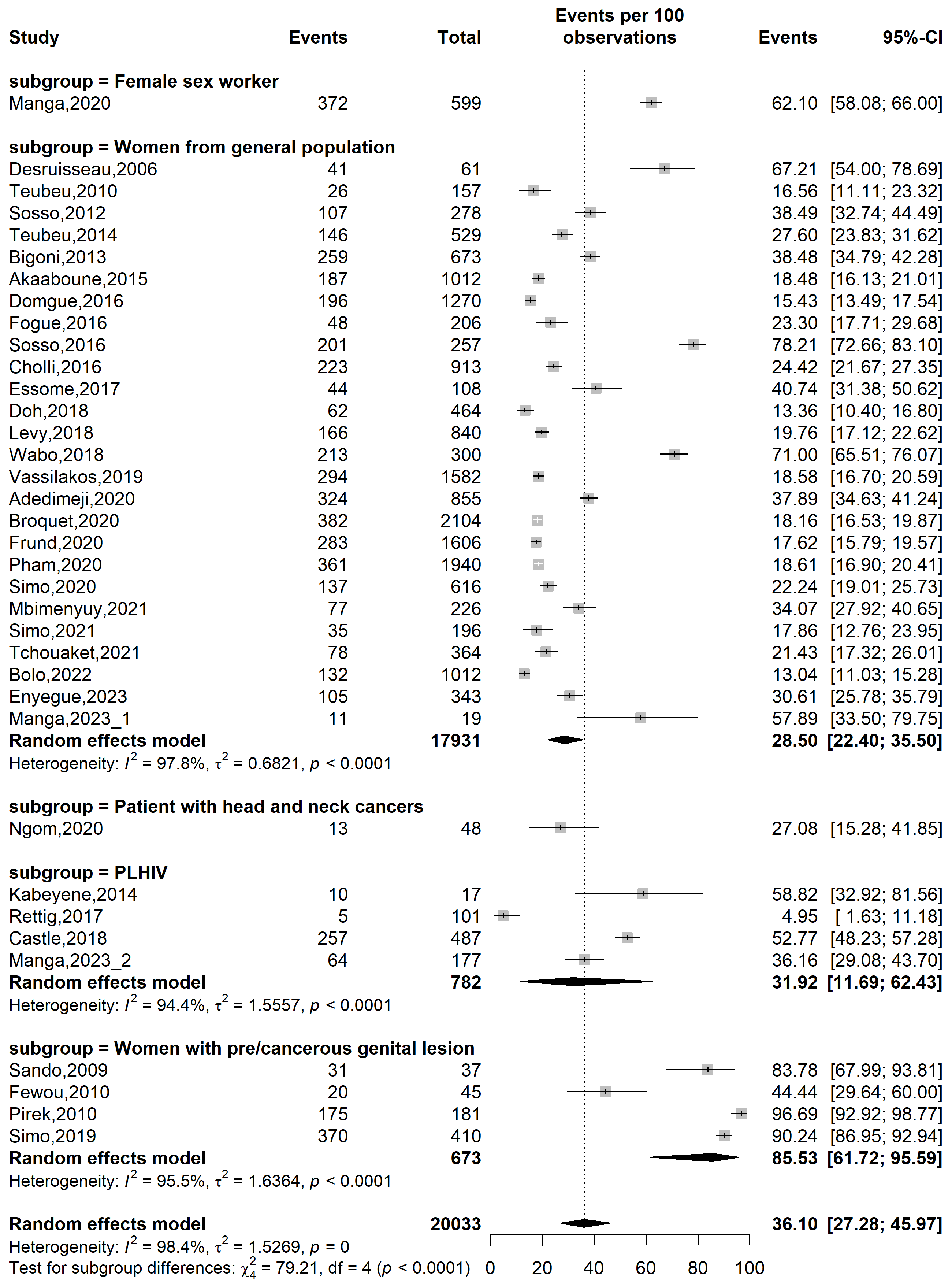

**Supplementary Fig. 7** Pooled HPV prevalence in Cameroon by type of participants

**Risk Factor**

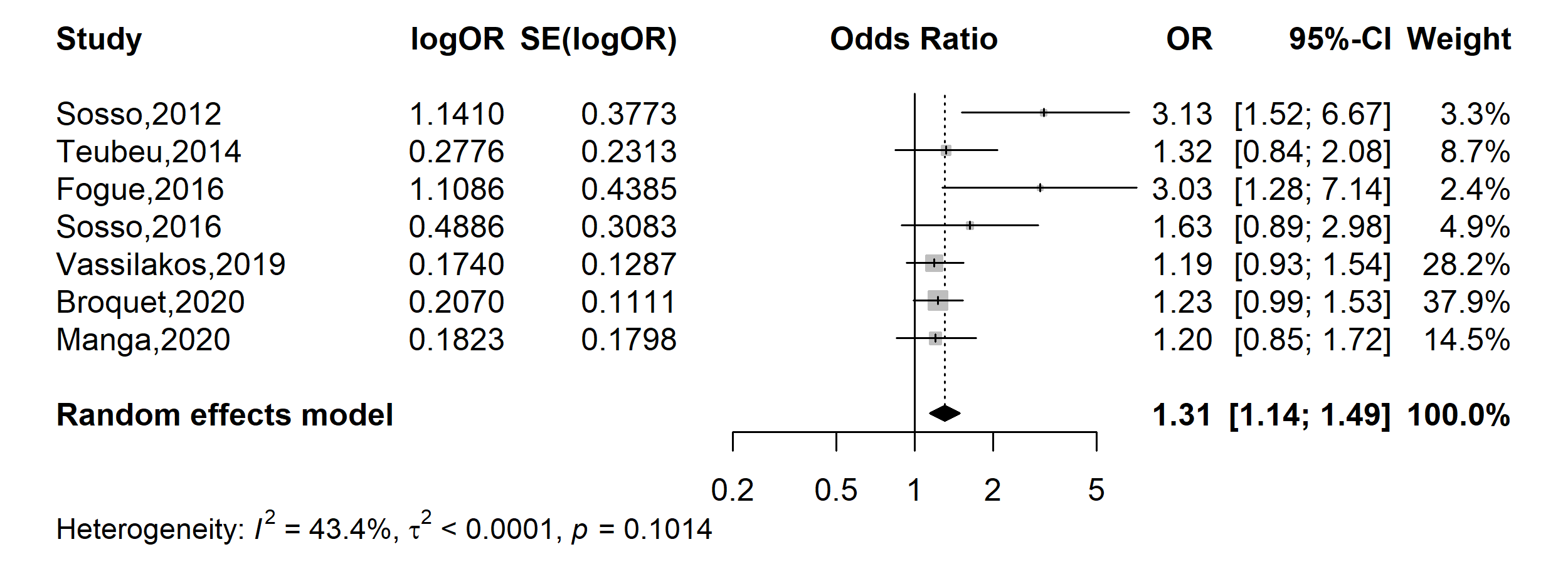

**Supplementary Fig. 8** Pooled odds ratio of HPV positivity in Cameroon (Age: ˂40 vs. 40+ years)

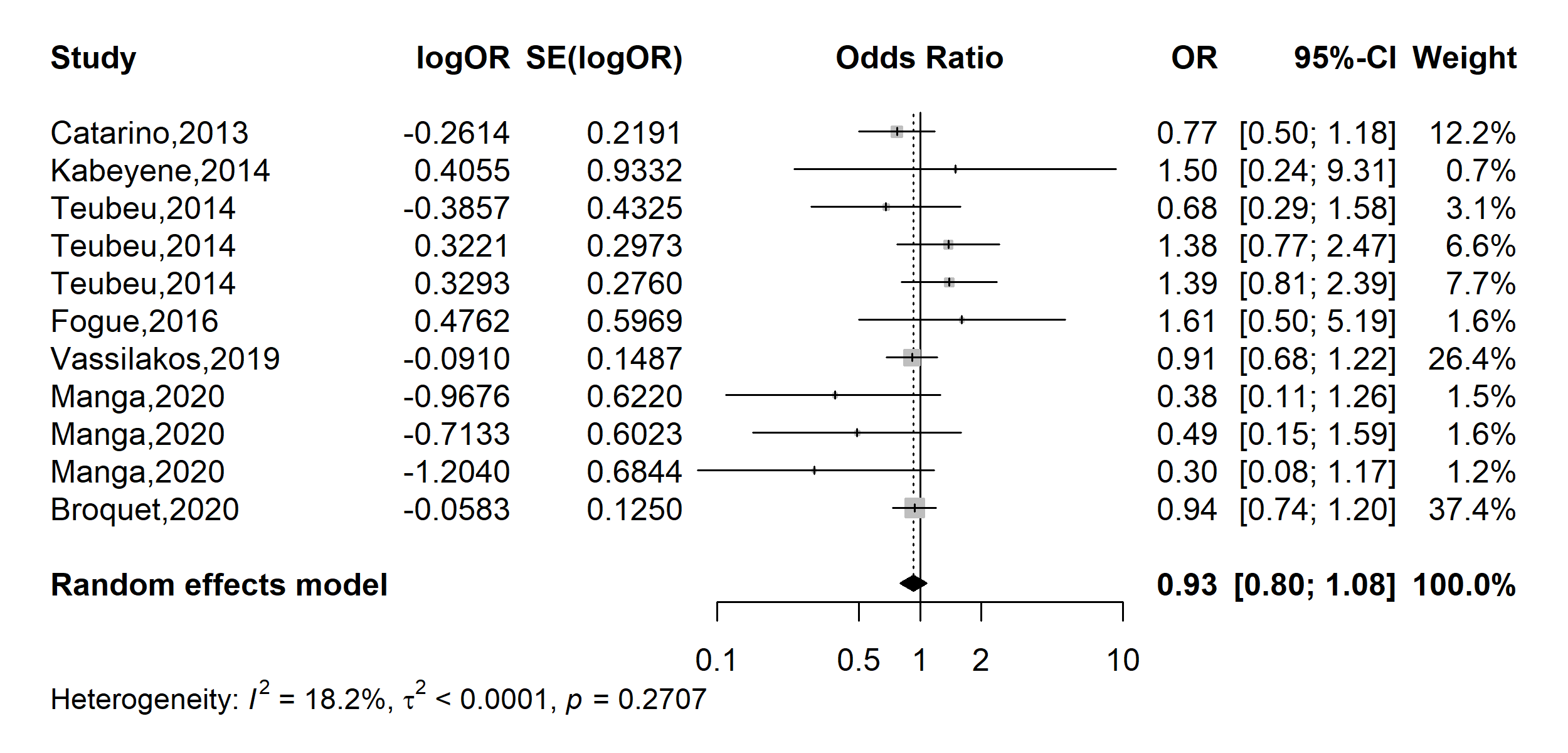

**Supplementary Fig. 9** Pooled odds ratio of HPV positivity in Cameroon (Educational level: Tertiary vs. Other)

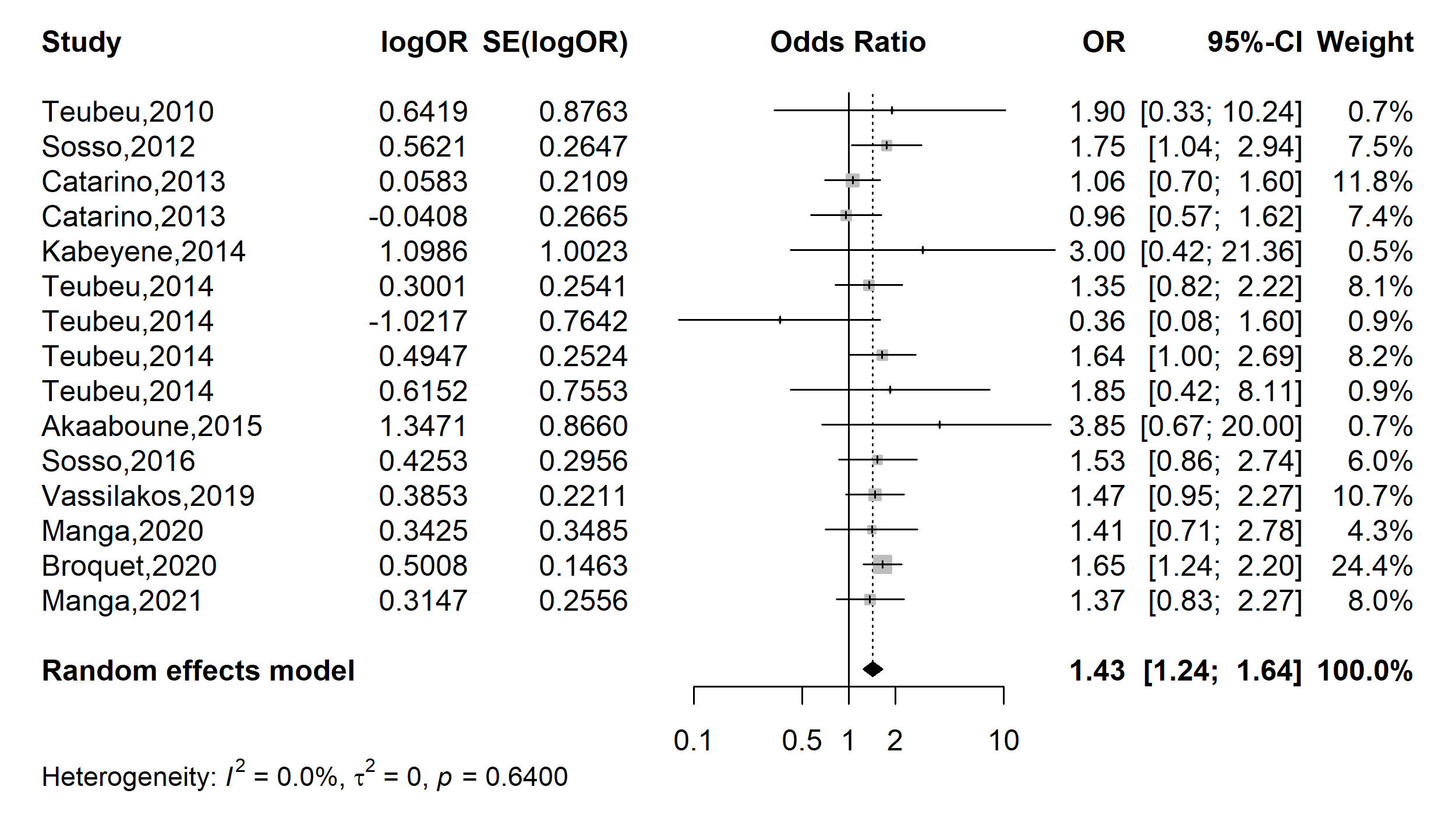

**Supplementary Fig. 10** Pooled odds ratio of HPV positivity in Cameroon (Marital status: Unmarried vs. In partnership)

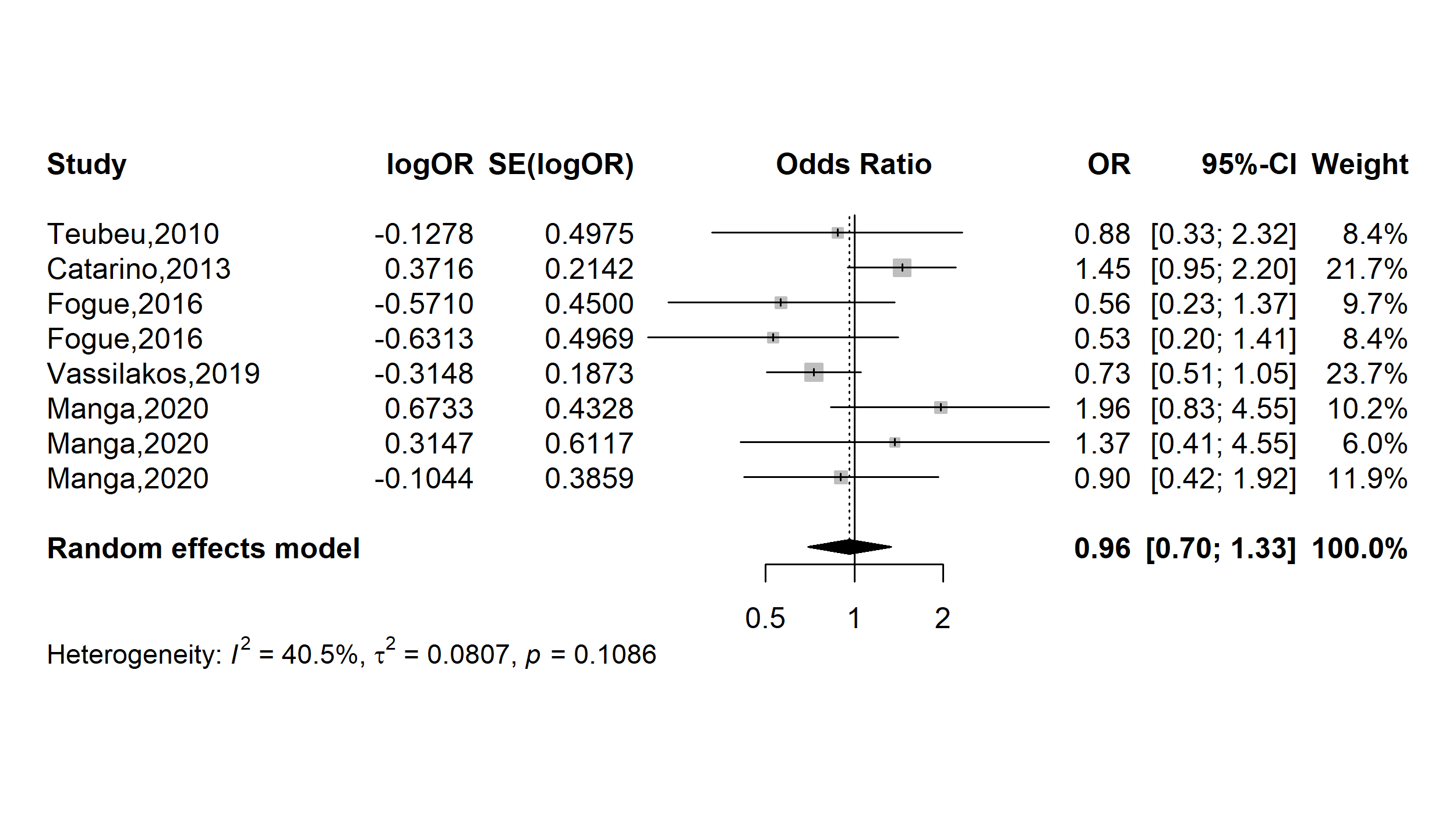

**Supplementary Fig. 11** Pooled odds ratio of HPV positivity in Cameroon (Occupation: Housewife vs. Other occupation)

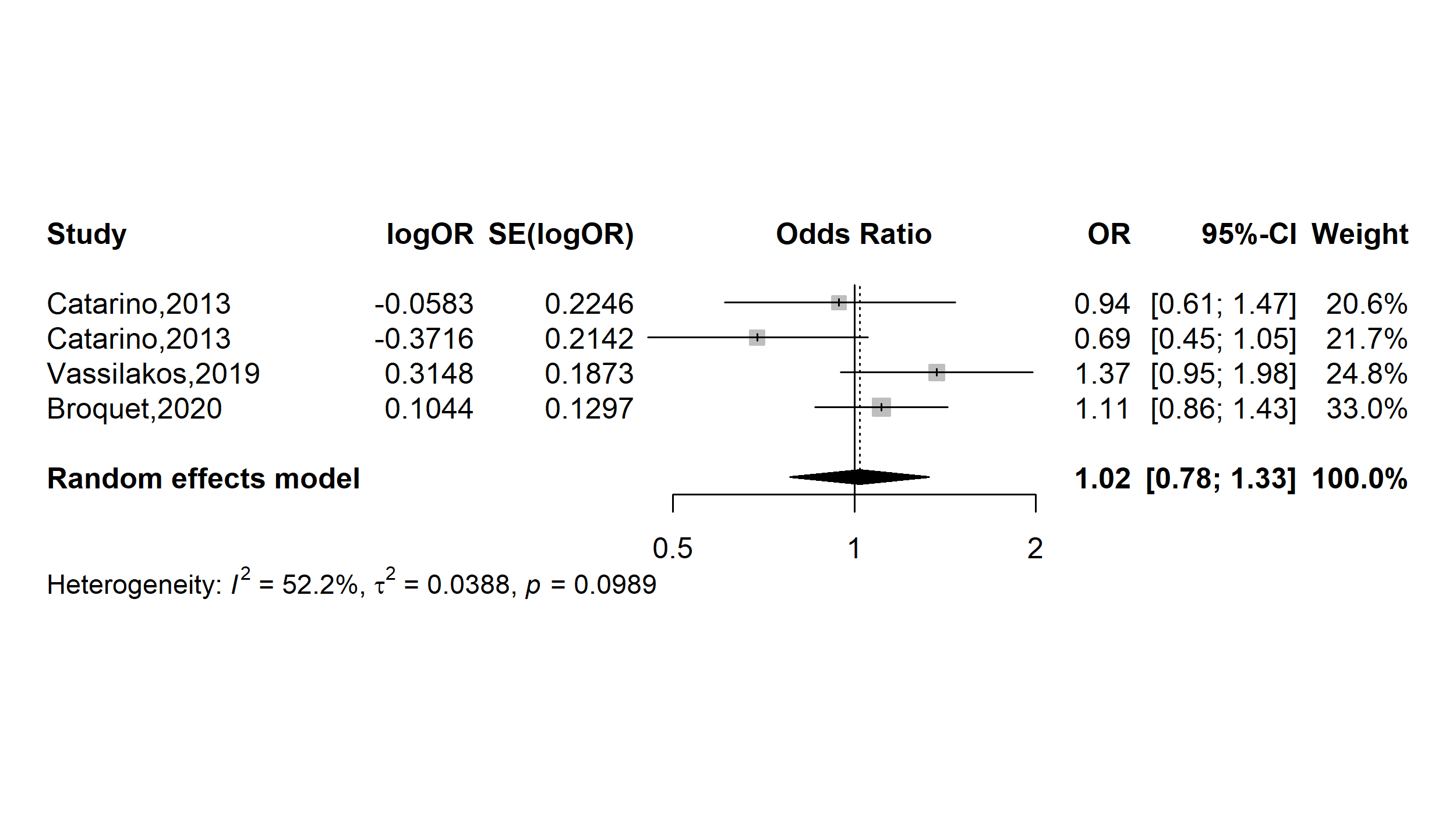

**Supplementary Fig. 12** Pooled odds ratio of HPV positivity in Cameroon (Occupation: Employed vs. Other occupation)

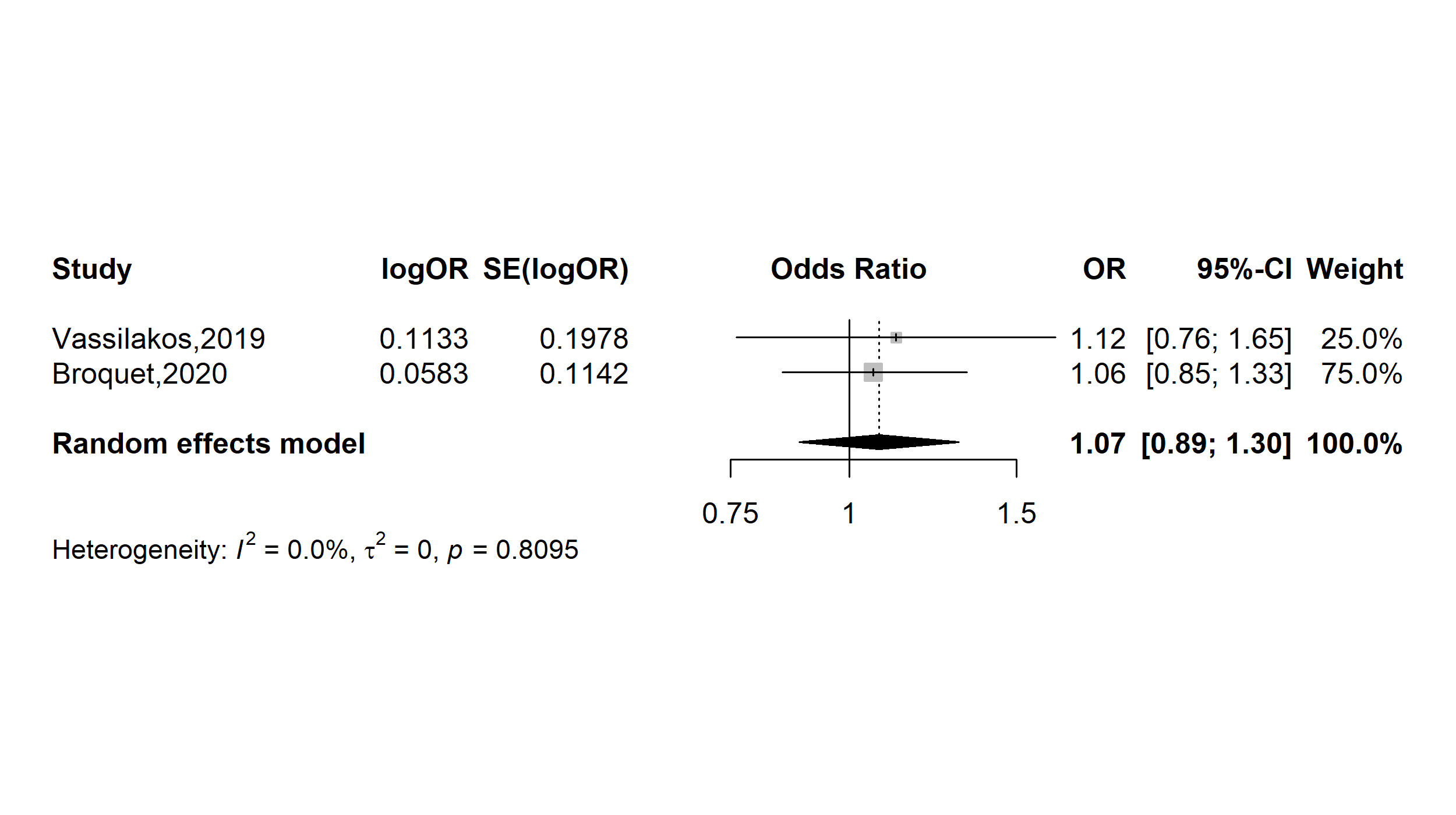

**Supplementary Fig. 13** Pooled odds ratio of HPV positivity in Cameroon (Menarche: ˂13 vs. 13+ years)

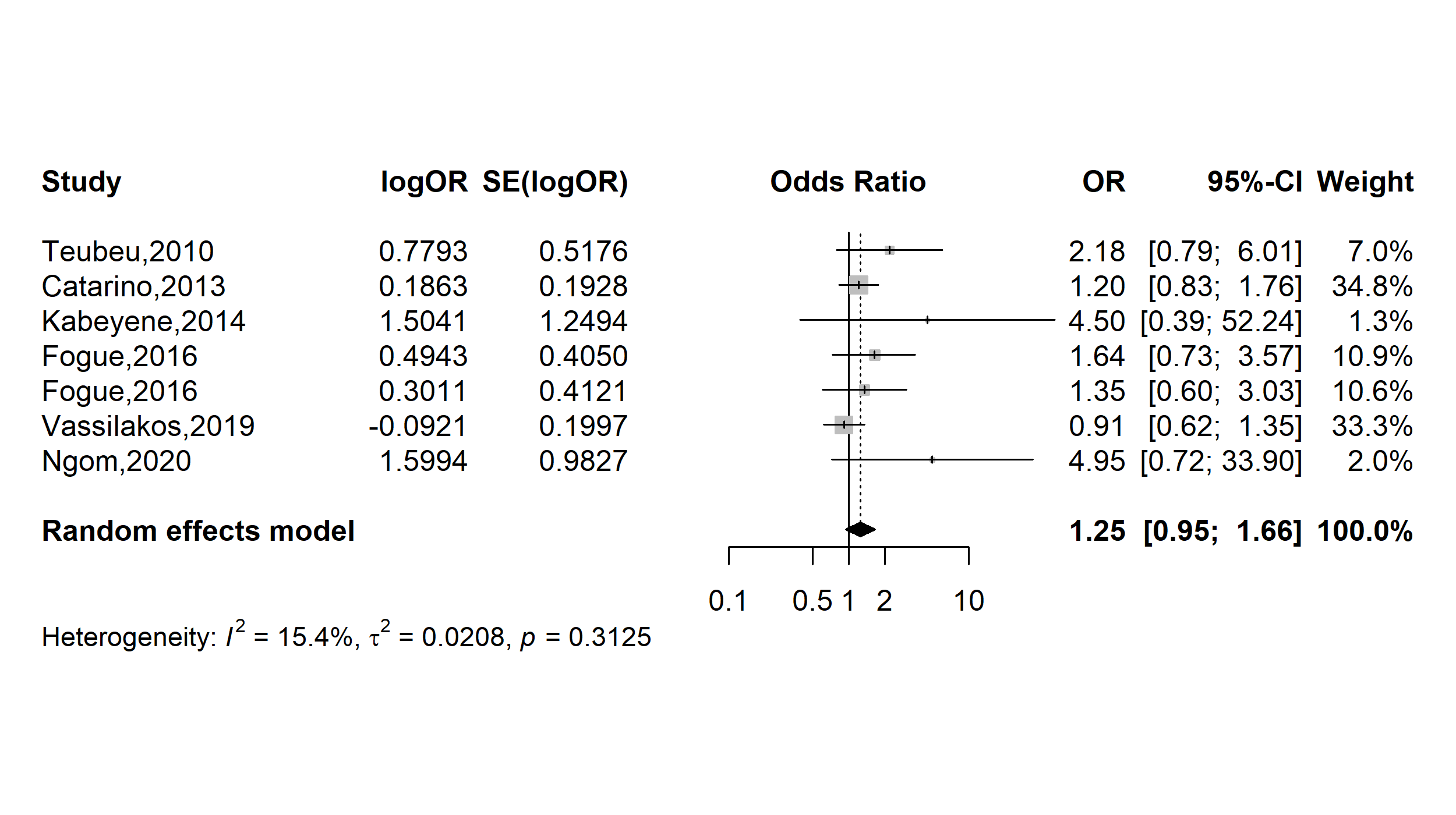

**Supplementary Fig. 14** Pooled odds ratio of HPV positivity in Cameroon (Age first sexual intercourse: ˂16 vs. 16+ years)

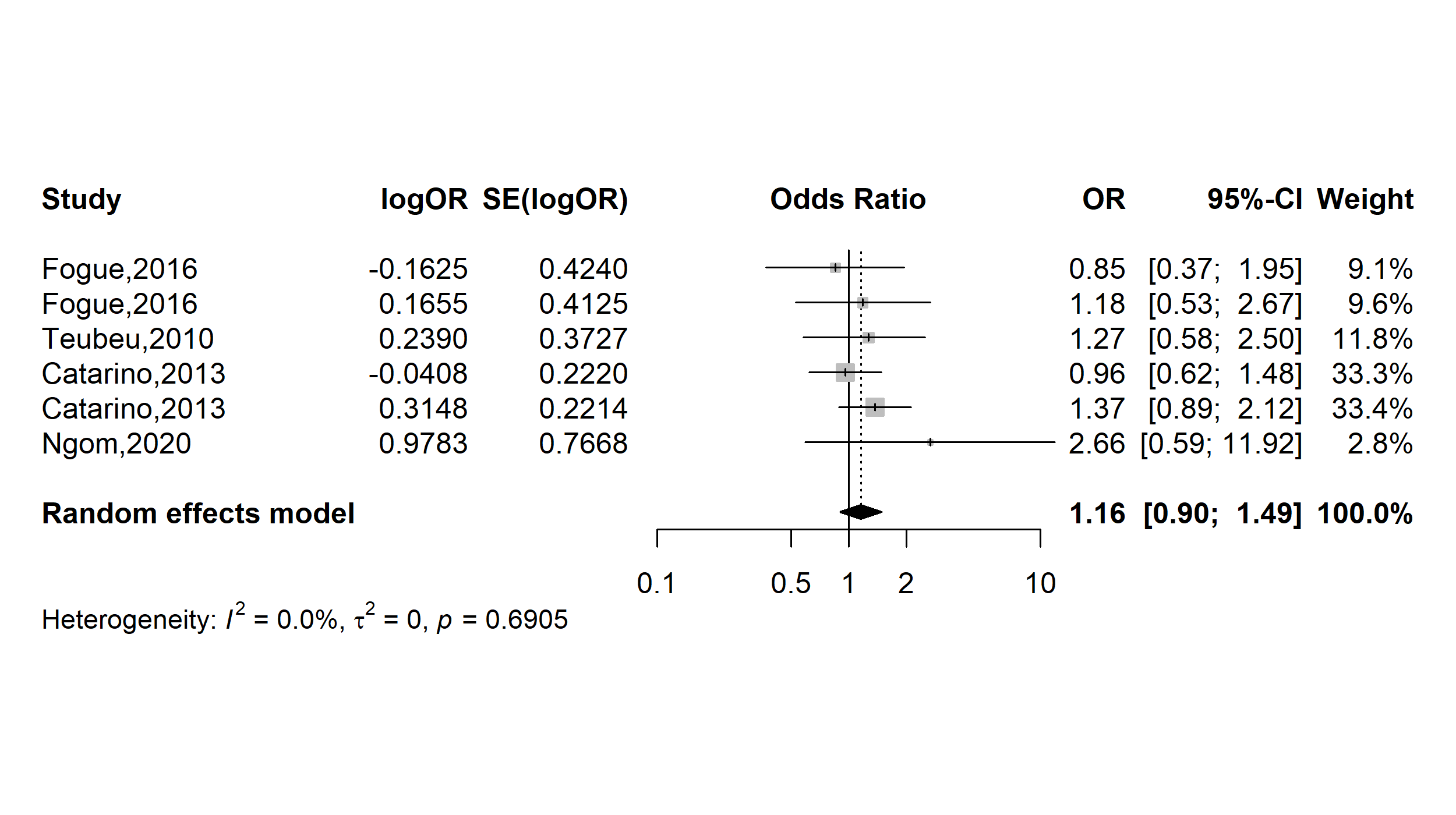

**Supplementary Fig. 15** Pooled odds ratio of HPV positivity in Cameroon (Number of sexual partners: 2+ vs. ˂2)

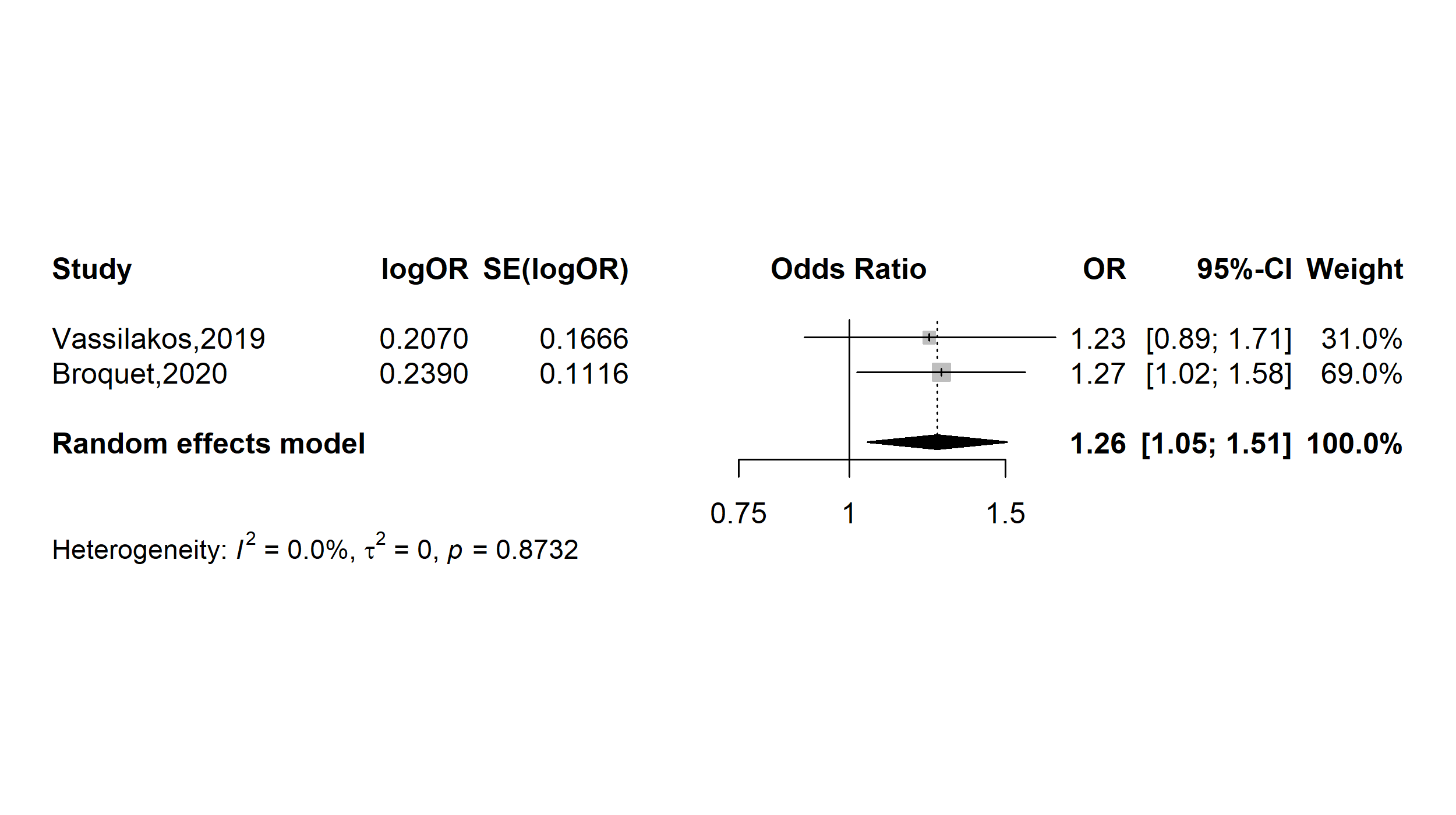

**Supplementary Fig. 16** Pooled odds ratio of HPV positivity in Cameroon (Number of sexual partners: 5+ vs. ˂5)

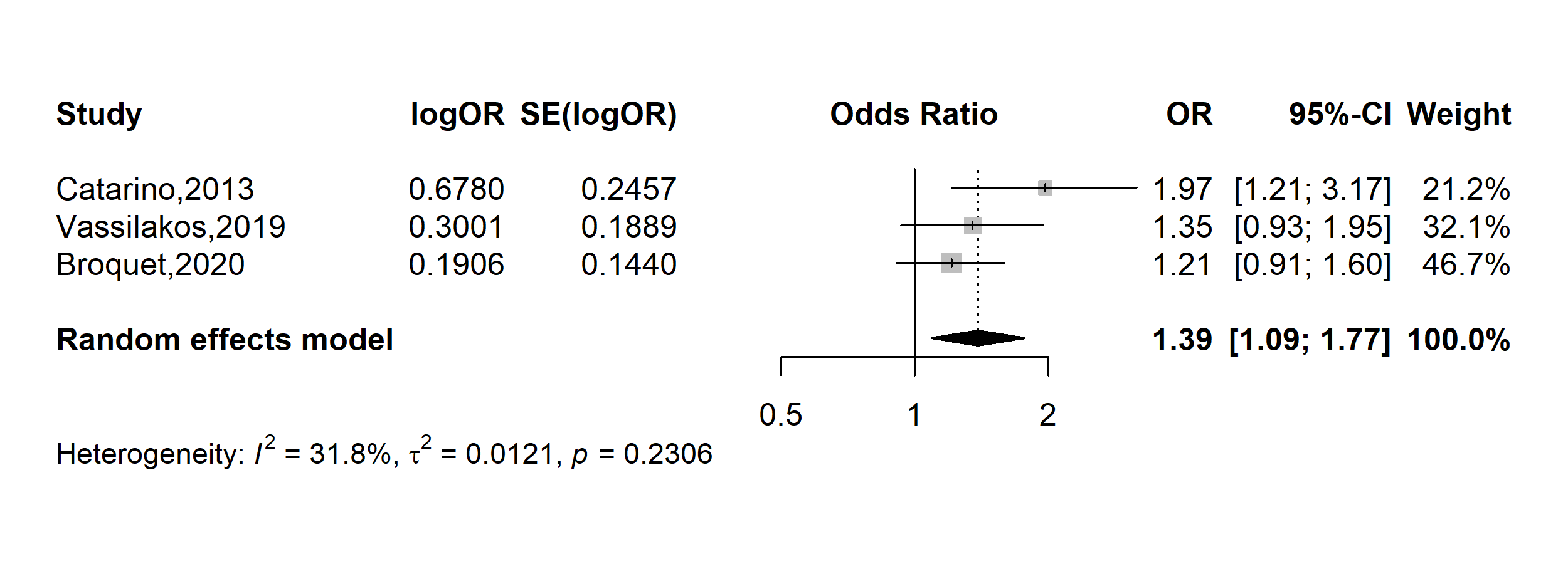

**Supplementary Fig. 17** Pooled odds ratio of HPV positivity in Cameroon (Contraception: Hormonal vs. None)

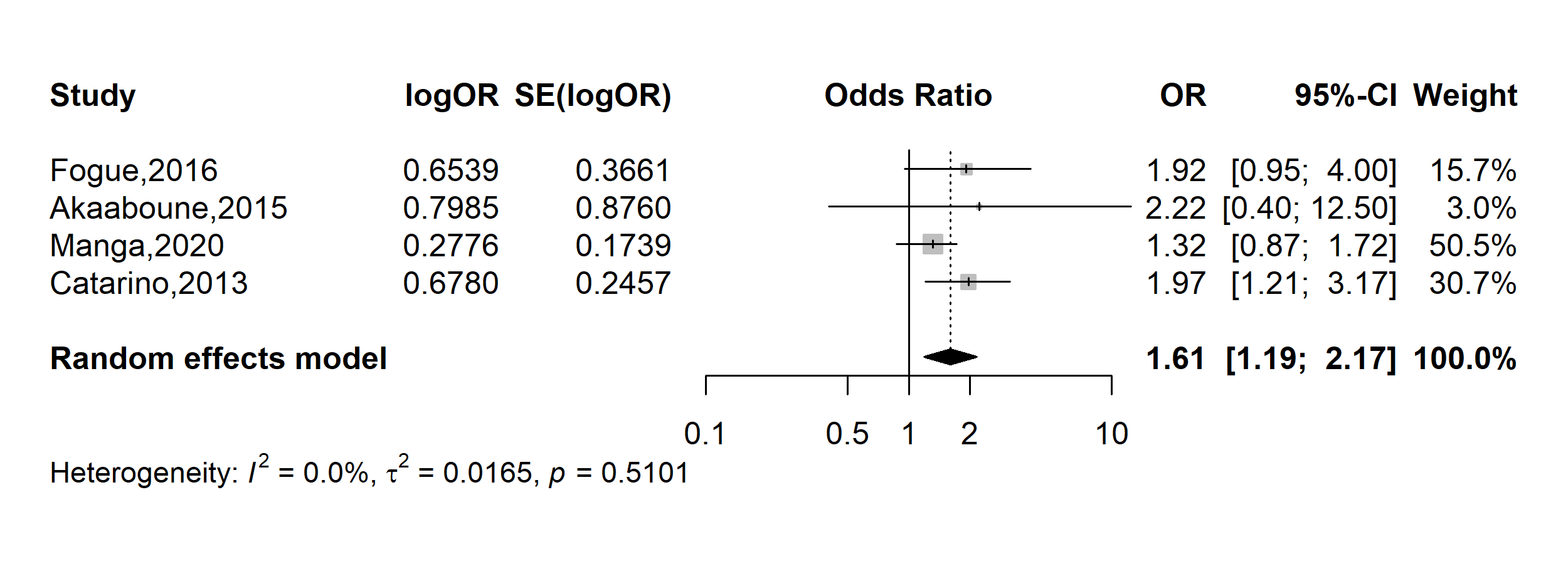

**Supplementary Fig. 18** Pooled odds ratio of HPV positivity in Cameroon (Contraception: Condom vs. None)

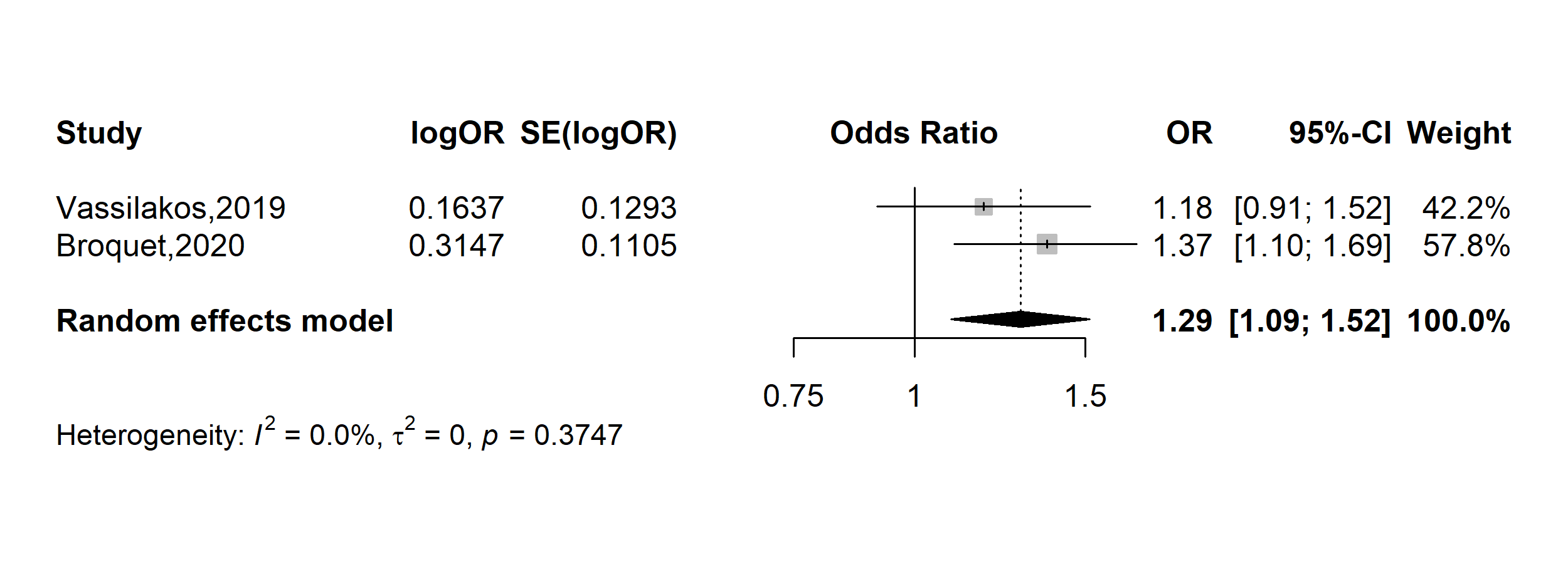

**Supplementary Fig. 19** Pooled odds ratio of HPV positivity in Cameroon (parity: ˂4 vs. 4+)

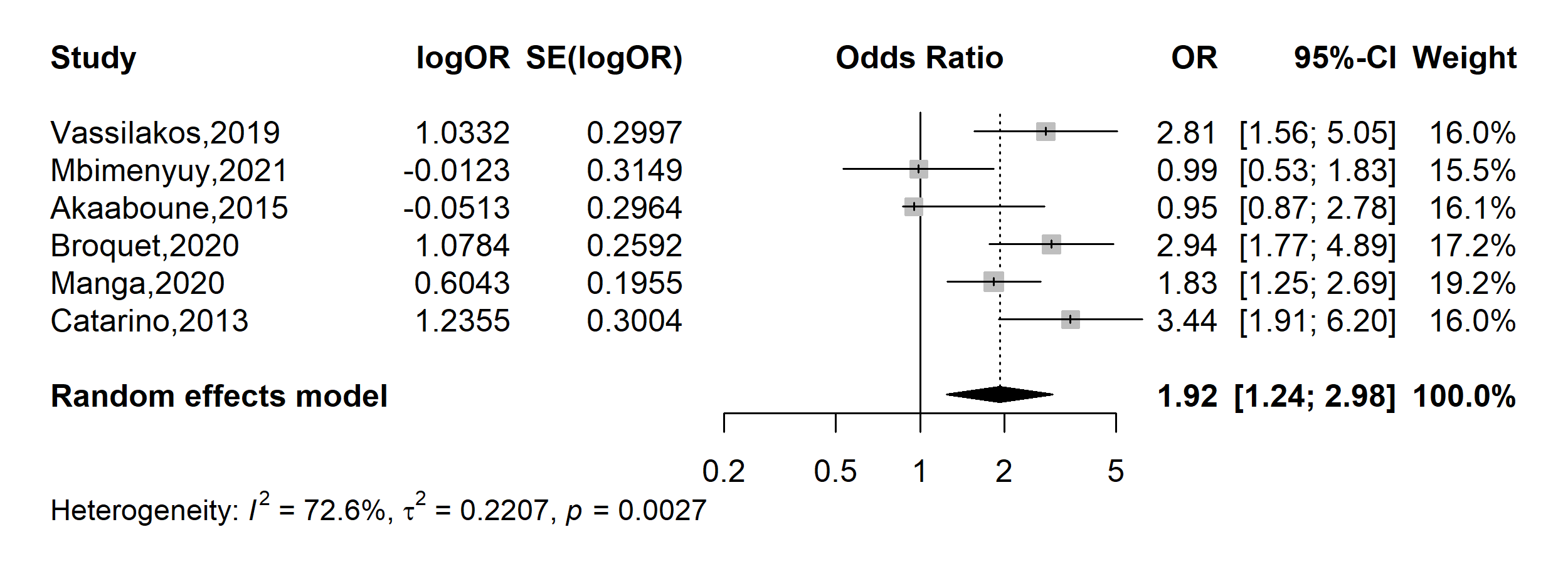

**Supplementary Fig. 20** Pooled odds ratio of HPV positivity in Cameroon (HIV status: positive vs. negative)

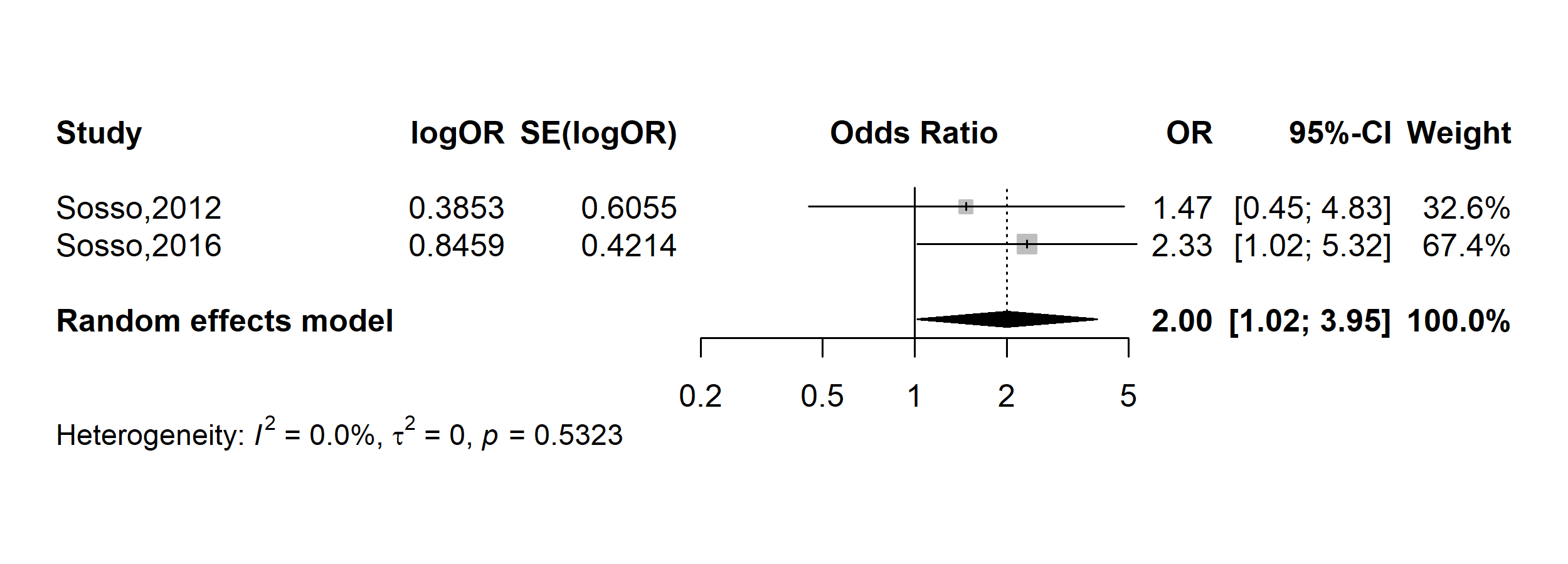

**Supplementary Fig. 21** Pooled odds ratio of HPV positivity in Cameroon (CD4 count in cells/mm^3^: ˂500 vs. 500+)

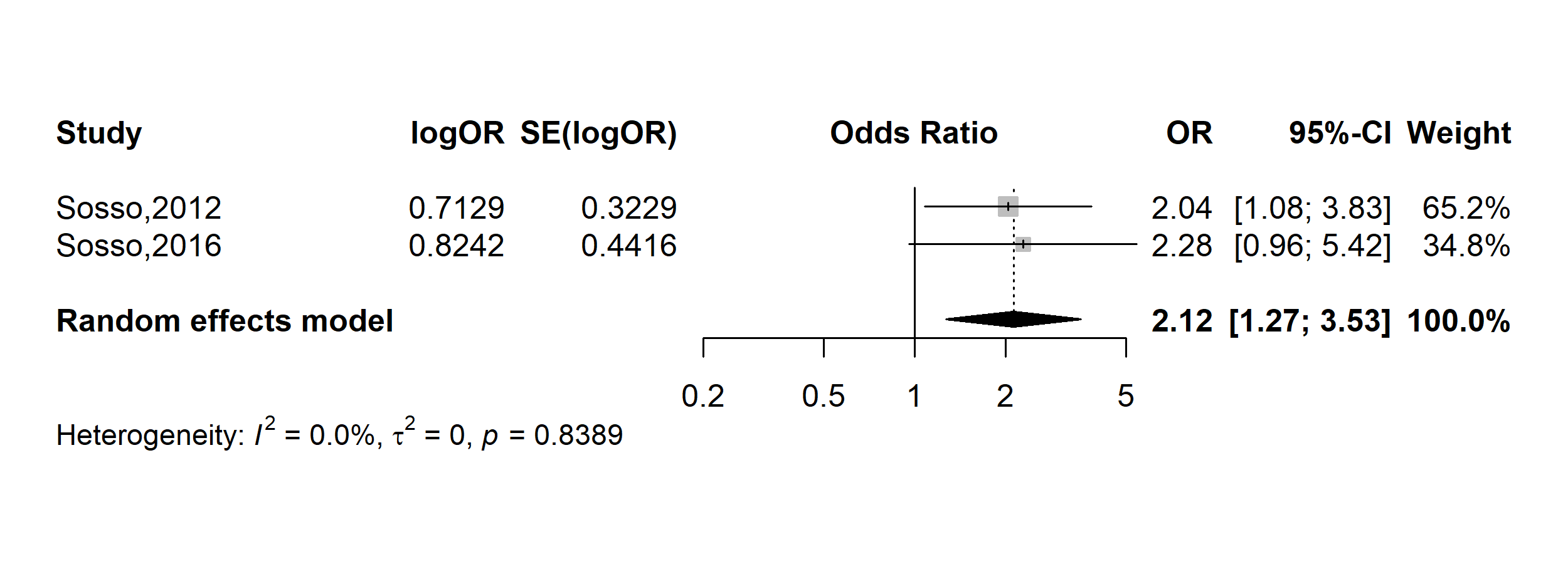

**Supplementary Fig. 22** Pooled odds ratio of HPV positivity in Cameroon (Viral load in copies/mL: ˂1000 vs. 1000+)
